# A cost-effectiveness analysis of Molnupiravir and Paxlovid in three African countries

**DOI:** 10.1101/2023.07.05.23292205

**Authors:** Ijeoma Edoka, Tom Drake, Peter Baker, Nicaise Ndembi, Raji Tajudeen, Elias Asfaw, Javier Guzman, Justice Nonvignon, Jean Kaseya

## Abstract

**Objective:** To assess the cost-effectiveness of two COVID-19 oral antivirals (COAVs) Paxlovid and Molnupiravir compared to the standard of care, in Ghana, Rwanda and Zambia.

**Methods:** We modelled costs (2022 US$) and health outcomes in the acute phase of the COVID-19 disease from a public payer’s perspective in three unvaccinated target populations – (1) patients aged 65 years and above (elderly); (2) adult patients with at least one other underlying risk factors for disease severity; and (3) all adult patients. In addition, we conducted a series of sensitivity and scenario analyses.

**Results:** In elderly patients, Paxlovid was less costly and more effective (i.e., dominated) than standard of care in all three study countries. Molnupiravir dominated standard of care in Rwanda and Zambia and an incremental cost-effectiveness ratio (ICER) was estimated at US$1023.58 per disability-adjusted life year (DALY) averted in Ghana. In adults with other underlying risk factors, Paxlovid dominated in Rwanda and Zambia while Molnupiravir dominated in Rwanda. Neither Paxlovid nor Molnupiravir were cost-effective in the all-adult group in any country context. Incremental net monetary benefit for Paxlovid was consistently higher than for Molnupiravir. In COVID-19 vaccinated patients, Paxlovid was cost-effective for elderly patients in Zambia and Rwanda but not in Ghana. Key determinants of cost-effectiveness were COAV price, likelihood of early treatment initiation, and hospitalization rates.

**Conclusion:** In African settings similar to Zambia, Ghana or Rwanda, COAVs could be cost-effective in populations who are unvaccinated, and at high risk of progression to severe COVID-19. More evidence is needed to determine cost-effectiveness for patients that are unvaccinated but have previously been infected with COVID-19 and may have developed some immune protection.

**Key messages:**

- What is already known on this topic – Two COVID-19 oral antivirals (COAVs), Molnupiravir and Paxlovid have been shown to represent good value for money in high-income countries. However, there is a dearth of evidence on the cost-effectiveness of these drugs in African countries.
- What this study adds-This study finds that COAVs are likely to be cost-effective in populations who are unvaccinated and at high risk of progression to severe COVID-19. However, the probability of Molnupiravir being cost-effective was consistently lower than the probability of Paxlovid being cost-effective. Early treatment initiation, COAV price and baseline hospitalization rates had the largest impact on the cost-effectiveness of both COAVs in unvaccinated patients. More evidence is needed on the cost-effectiveness for patients who have previously been infected with COVID-19.
- How this study might affect research, practice, or policy – This study broadly supports African governments decisions to not procure substantial quantities of either COAV but is evidence that Paxlovid could be good value for money when treating very specific populations.

## Introduction

COVID-19 has had a devastating impact in many African countries, both through its direct effect on the lives and livelihoods of affected populations and, indirectly, through its impact on health systems and the redirection of resources away from other essential services ^1, 2^.

The global roll-out of COVID-19 vaccines has contributed significantly to reducing the health and economic impact of COVID-19. However, disparities in COVID-19 vaccine coverage between high-income and low– and middle-income countries (HICs and LMICs) have highlighted global failures in achieving equitable distribution and delivery^3^. Since 2022, therapeutic interventions for the clinical management of COVID-19 both in the outpatient and inpatient setting have become available ^4^, offering more options in the fight against the disease. Two COVID-19 oral antivirals (COAVs), Pfizer’s nirmatrelvir and ritonavir (Paxlovid) and Merck’s Molnupiravir have received Emergency Used Authorization (EUA) for use in several high income countries and have been recommended for use in African Union Member states by the Africa Centres for Disease Control and Prevention^5^. Initiatives such as the voluntary licensing agreement between Pfizer, Merck and the Medicines Patent Pool and the test and treat programs announced by USAID and the COVID Treatment Quick Start Consortium in 2022 have made available low priced, generic versions of patent-protected medicines and donations for LMIC use. When compared to placebos, these COAVs have been shown to reduce the risk of hospitalizations and deaths in unvaccinated outpatients with at least one risk factor for progression to more severe conditions ^6, 7^. Given the low levels of vaccination coverage in Africa, the availability and use of these therapeutic agents could be important for minimizing the impact of COVID-19 on individuals and on fragile health systems in Africa. Initiatives such as the Accord initiative by Pfizer and the voluntary licensing agreement between Pfizer, Merck and the Medicines Patent Pool could see the manufacture of affordable generic versions of patent-protected medicines on a not-for-profit basis in several LMICs at prices significantly lower than is offered to high income countries^8, 9^.

As more COAVs become available, decision-makers in LMICs will need to weigh various factors, including clinical impact, cost, availability, and feasibility of use, in deciding which COAV to recommend for use within their context. Given growing budget constraints facing many LMICs, not least due to the COVID-19 pandemic, an understanding of the value-for-money of COAVs is likely to be crucial to informing policy decisions on the provision of COAVs either to entire populations affected by COVID-19 or to specific sub-populations. Some evidence suggests that Paxlovid and Molnupiravir can represent good value for money in some contexts ^10–12^. However, these studies largely focus on populations in high-income countries and, given differences between settings along dimensions that can influence results of cost-effectiveness analyses, these findings may not be transferable to LMICs. For example, the burden of disease (hospitalizations and deaths) in Africa has been markedly lower compared to other regions, which could result in less favorable cost-effectiveness outcomes. On the other hand, the higher probability of death following hospitalization in Africa due to poor health systems ^13^ may mean that the value of COAVs in terms of reducing the need for hospitalization, may be greater in the Africa setting. To be effective, COAV treatment must be initiated within five days of symptom onset ^6, 7, 14^. Therefore, the lower likelihood of early treatment initiation may limit the effectiveness of COAVs (Caraco et al., 2022; Hammond et al., 2022; Jayk Bernal et al., 2022) in LMICs where COVID-19 testing rates and testing policies may not be as rigorous as in high income countries to allow early diagnosis and treatment initiation. Cost-effectiveness analyses that consider the nuances within the African context is crucial for understanding the conditions under which COAVs would represent good value for money for African countries.

## Methods

This study assesses the cost-effectiveness of Paxlovid and Molnupiravir in unvaccinated adult populations, including sub-populations at high risk of disease progression in three African countries. The study countries were selected to reflect regional distribution of countries within the Africa Union and represent a range of factors that may directly or indirectly impact the effectiveness and thus, the cost-effectiveness of COAVs. These include the probability of early treatment initiation and COVID-19 vaccination coverage. Countries were stratified by vaccination coverage and COVID-19 test rates (a proxy for early diagnosis and treatment) ^15^ and representative countries –Ghana, Rwanda, and Zambia—selected from each stratum (Figure S1, Table S1).

The cost-effectiveness analysis was conducted from a public healthcare payer’s perspective by comparing the costs and health outcomes of a 5-day course of Paxlovid (Nirmatrelvir 300mg+Ritonavir 100mg twice daily) and Molnupiravir (800mg twice daily) to usual care (when no COAV treatment is administered). A decision tree was constructed in Microsoft Excel, to model adult patients with mild/moderate symptoms of COVID-19 treated in the outpatient setting. The model estimated health outcomes and costs in three target populations-(1) all unvaccinated adult patients with a starting age equivalent to the average age of COVID-19 in the adult population of each study country; (2) unvaccinated patients aged 65 years and above; and (3) unvaccinated adult patients with at least one other underlying risk factors for COVID-19 disease severity.

We modeled disease progression in the acute phase of the disease (30 days). Patients enter the model either remaining in the outpatient setting or progressing to hospitalization (Figure 1). Those remaining in the outpatient setting either survive or die from other causes. For those hospitalized, we simulated different levels of care depending on disease severity –mild/moderate, severe, and critical—and their corresponding survival rates. Given the need for treatment initiation within five days of symptom onset, we explicitly modeled the probability of early initiation using country-specific COVID-19 test rates as proxies. We assumed no treatment effect for those initiating treatment after five days of symptom onset^6, 7, 14^.

**Figure 1:**
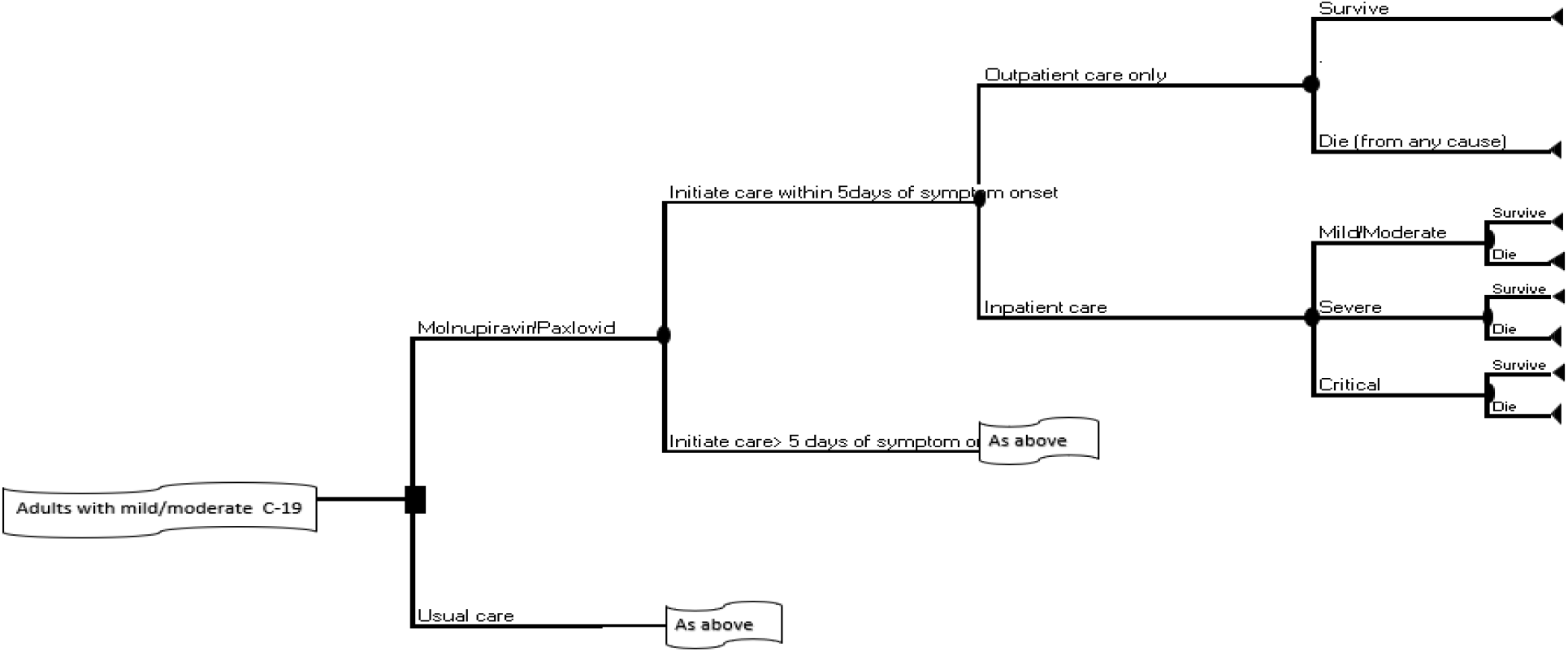
Diagrammatic representation of the model.

Both incremental cost-effectiveness ratios (ICERs) and incremental net monetary benefits (INMBs) were estimated. Cost-effectiveness was determined using country-specific cost-effectiveness threshold that reflects the estimated marginal productivity of each study country’s health system ^16^ (Table 1).

**Table 1:**
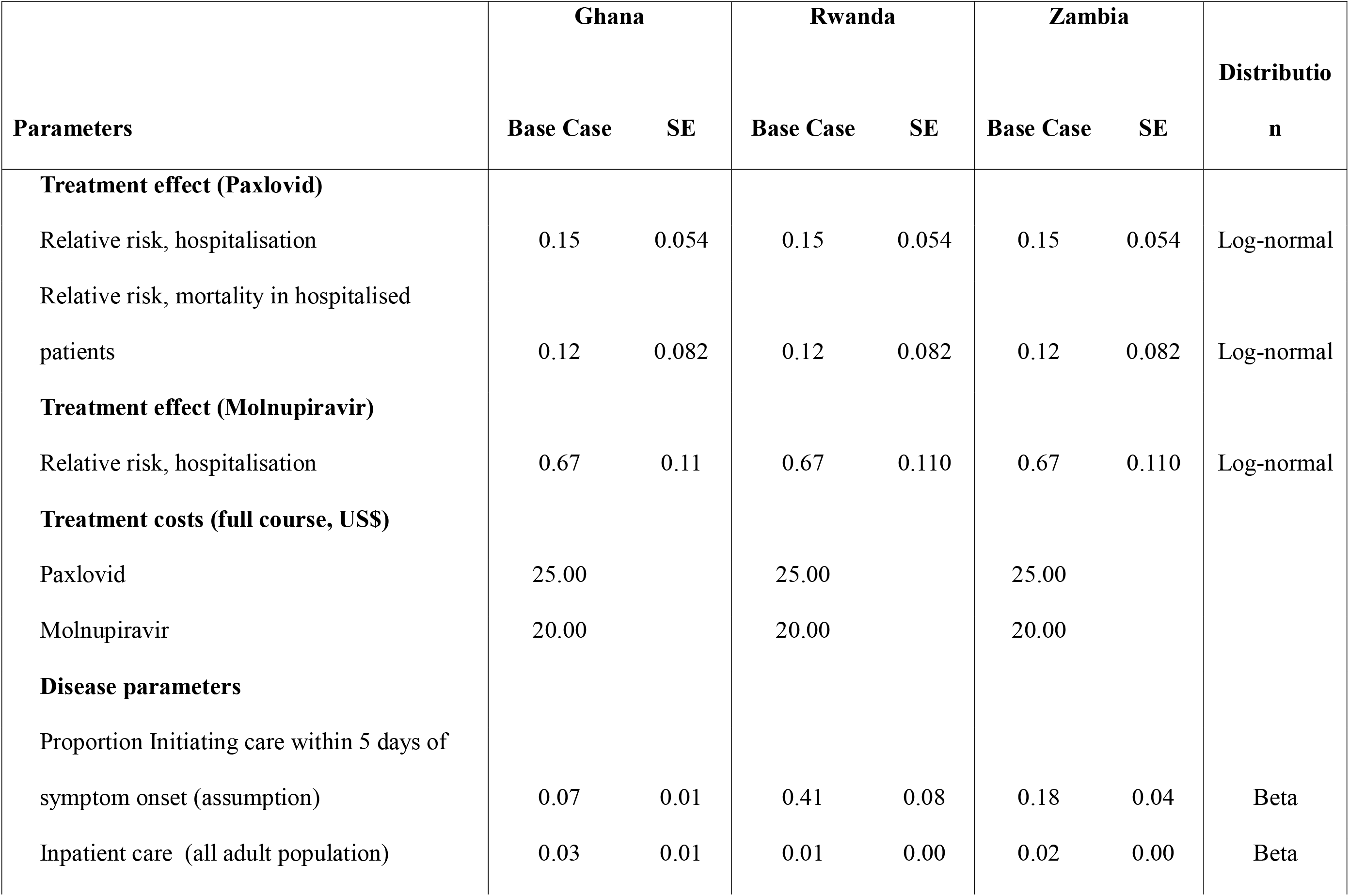
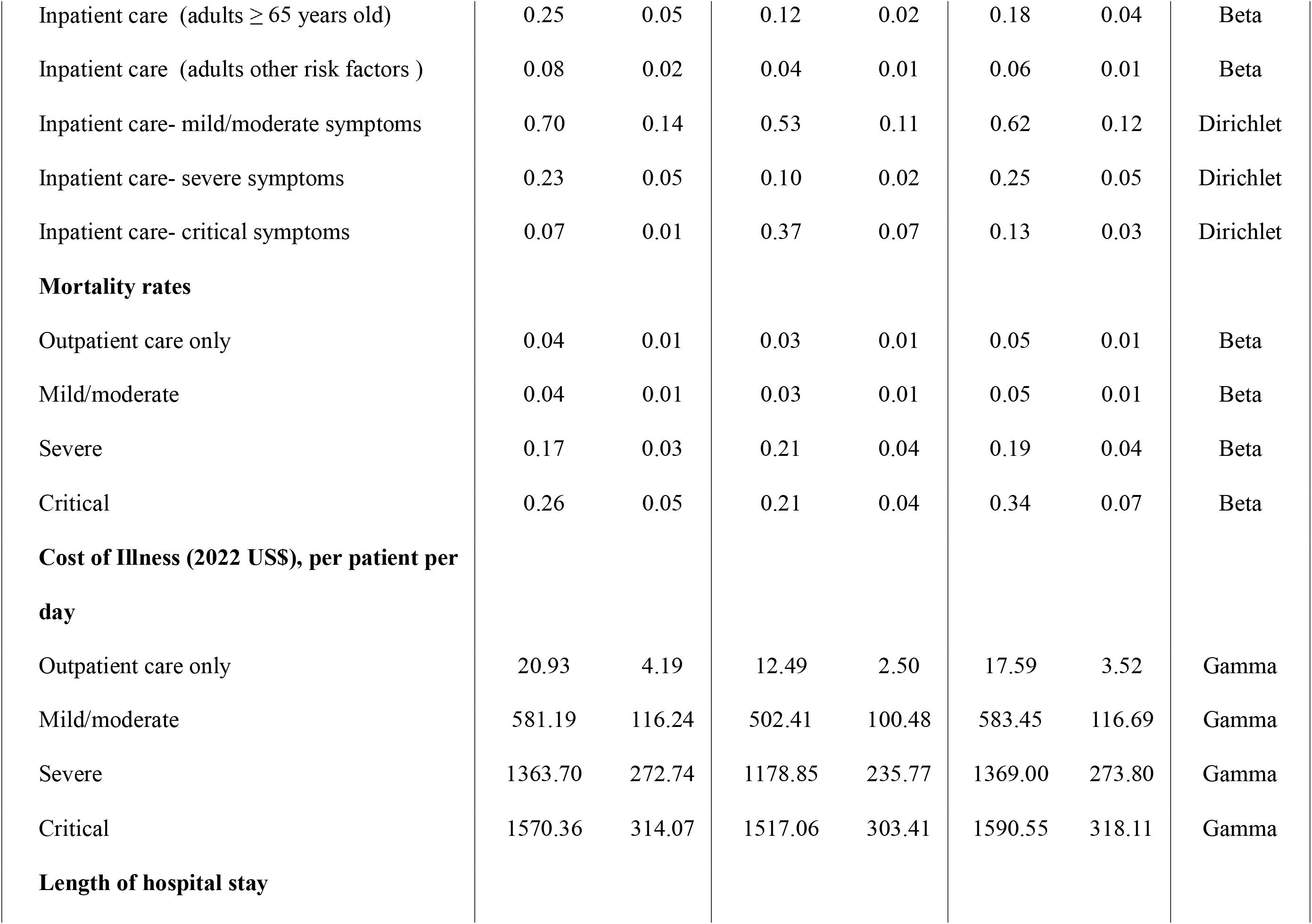
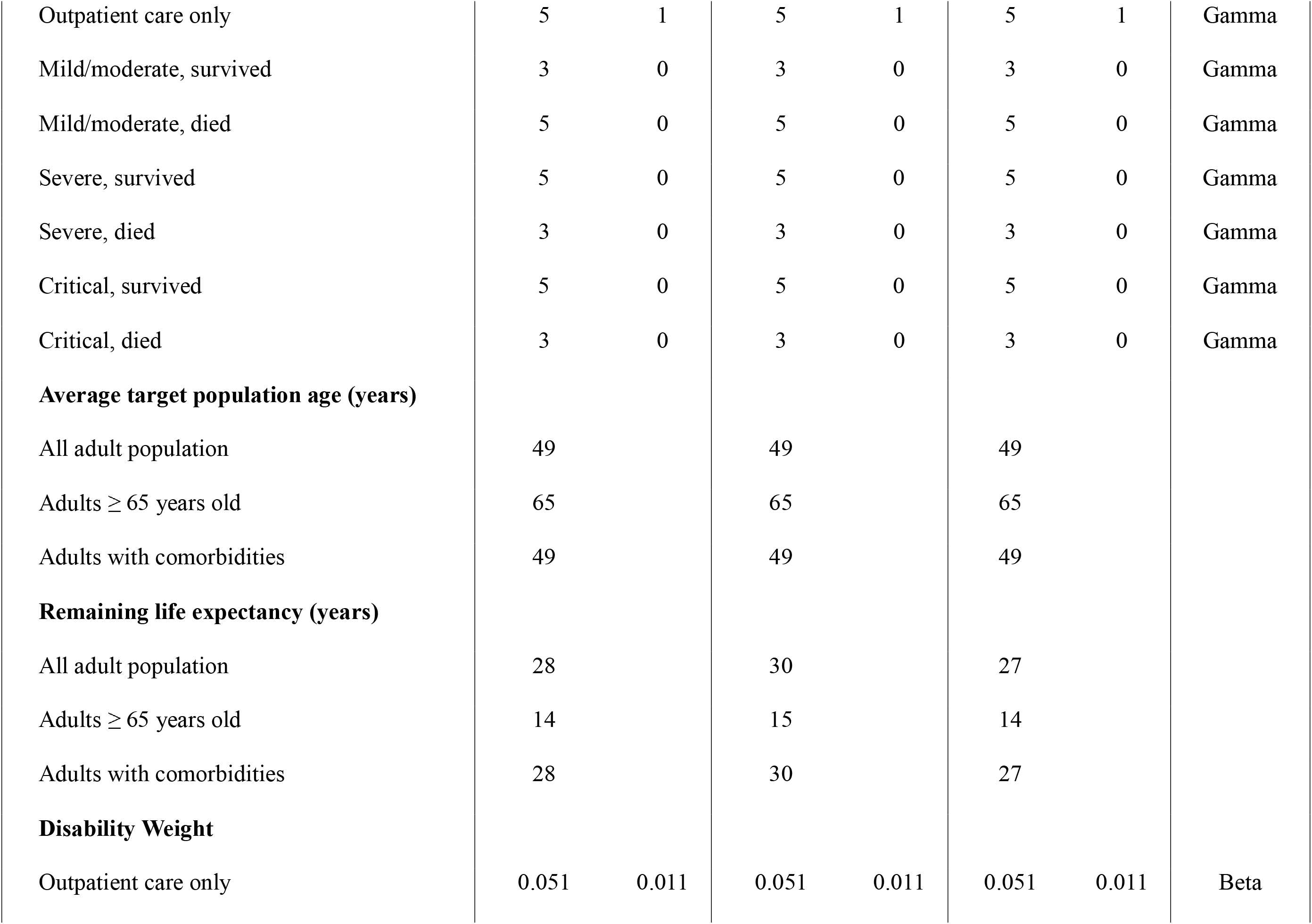
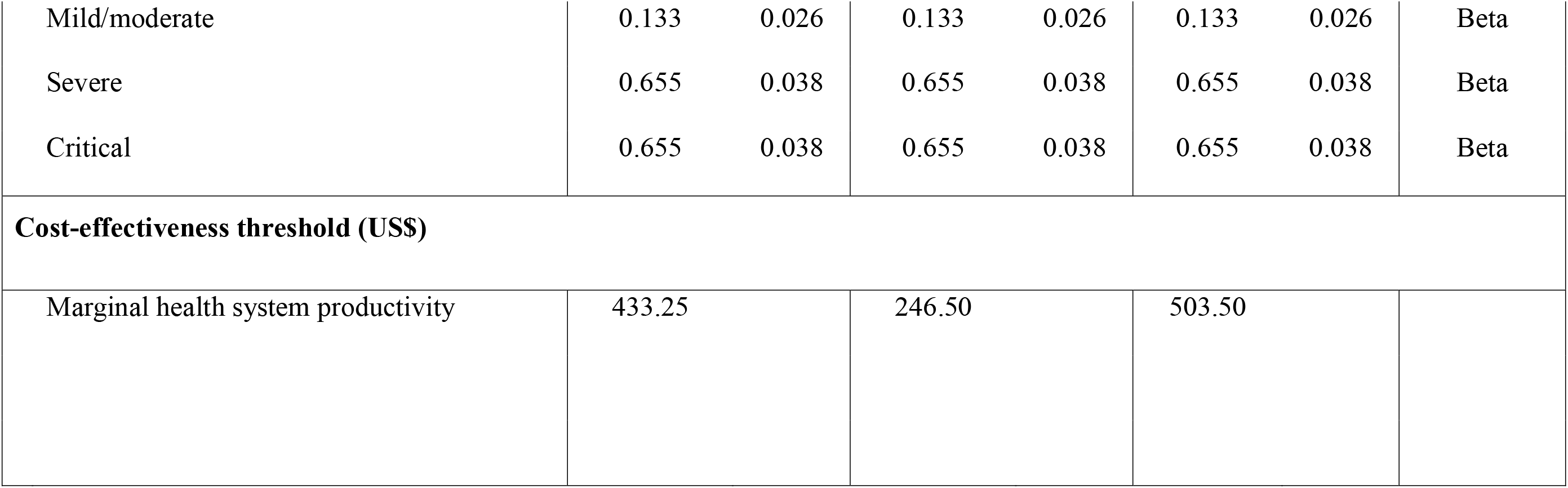
Model Parameters (Base case)

### Model Parameters

The model was populated using parameters obtained from various sources (Table 1): This includes:

### Disease parameters

For the adult population, country-specific disease parameters including hospitalization rates, in-hospital disease severity and survival rates, were obtained from publicly available sources or literature that reported on the epidemiological profile of the disease during the first phases of the pandemic ^13, 17–19^. Considering the current epidemiologic profile of the disease – potentially high natural immunity in Africa and the dominant milder COVID-19 Omicron variant ^20–22^, we adjusted hospitalization rates, inpatient disease severity and length of hospitalization downwards based on the relative disease severity between earlier COVID-19 variants and the Omicron variant ^20^. For older patients and patients with underlying risks factors, hospitalization rates was assumed to be 9 and 3 times, respectively, higher than hospitalization rates in the overall adult population ^23^.

### Clinical efficacy

The clinical benefits of Paxlovid and Molnupiravir were modeled as a reduction in the risk of hospitalization following initiation of COAV treatment within five days of symptom onset in unvaccinated patients ^6, 7^. In addition, we modeled a reduction in the risk of in-hospital mortality for Paxlovid but not for Molnupiravir given limited evidence on the efficacy of Molnupiravir on in-hospital mortality ^7^. COAV treatment effect was obtained from a meta-analysis of existing randomized control trials ^24^.

### Costs

Costs of clinical management of COVID-19 were obtained from existing literature. For Ghana, cost estimates were obtained from a primary costing study reporting context-specific estimates based on national treatment guidelines for outpatient– and inpatient management of COVID-19, disaggregated by disease severity ^25^. In the absence of context-specific cost estimates for Rwanda and Zambia, clinical management costs were obtained from Torres-Rueda et al^26^. Torres-Rueda et al^26^ estimated COVID-19 clinical management costs for 79 LMICs by extrapolating unit costs from three LMICs. However, a comparison to Ghana-specific cost estimates suggests that unit costs from Torres et al may have been underestimated. Therefore, clinical management costs for Rwanda and Zambia were scaled up using the proportionate difference between Torres-Rueda et al. ^26^ costs estimate for Ghana and the reported context-specific cost for Ghana.

A full 5-day course of Paxlovid was assumed to cost US$ 25 based on a maximum price per course negotiated by Clinton Health Access Initiative for the generic version of Paxlovid ^9^. A full course of Molnupiravir was assumed to costs US$20 based on pricing discounts offered LMICs through a voluntary licensing agreement between Merck, and the Medicines Patent Pool (MPP) to allow the manufacture of a cheaper generic version of Molnupiravir for LMICs ^8^. All costs were expressed in 2022 US$.

### Health outcomes

Health outcomes were expressed as disability-adjusted life year (DALYs) which was estimated as the sum of years of life lost (YLL) and years lived with disability (YLD). Remaining life expectancy at time of death for each country was obtained from the World Health Organization (WHO) life tables^27^. We used disability weights for each health state (mild/moderate, severe, and critical) based on the Global Burden of Disease Study 2019 (GBD 2019) estimates recommended by the European Burden of Disease Network Consensus COVID-19 model ^28^. YLL was discounted using a 5% discount rate based on assumptions of expected future economic growth in Africa ^29^.

### Sensitivity Analyses

To assess uncertainty in model inputs, deterministic and probabilistic sensitivity analyses (DSA and PSA) were conducted. The DSA varied individual parameters sequentially, over a specified range (Table S2) while holding all other parameters constant at their base case value.

PSA was conducted to assess the robustness of the results to uncertainty in model parameters, simultaneously. This was conducted by fitting appropriate distributions (Table 1) to each parameter ^30^ and running 1000 Monte Carlo simulations that drew parametric inputs from these distributions ^31^. Where available, standard errors of each parameter were used to determine the range but when not reported in data sources, standard errors were assumed to be 20% the mean value.

### Scenario Analyses

Scenario analyses were conducted to assess the impact of structural changes in the model on the results. Scenarios considered include (Table 2):

1. Cost-effectiveness in COVID-19 vaccinated patients (Scenario 1): Given that COVID-19 vaccination programmes have been implemented in our study countries, we assessed the cost-effectiveness of COAVs in vaccinated patients. However, recent evidence from a randomized control trial (RCT) suggests that Molnupiravir does not reduce the risk of hospitalization in vaccinated patients at high risk of disease progression^32^. While there is no reported RCT evidence on the efficacy of Paxlovid in vaccinated populations, a recent observational study suggests that Paxlovid effectively reduces the risk of hospitalization in vaccinated individuals^33^. Therefore, in this scenario, we assessed the cost-effectiveness of Paxlovid in vaccinated populations using effectiveness estimates from the observational study^33^. Furthermore, although the base case disease parameters of each country are likely to reflect both high natural immunity and COVID-19 vaccine effectiveness, we adjusted base case hospitalization rates downwards using vaccine efficacy and vaccination coverage of each study country ^15^. (Table 3).
2. 100% probability of early treatment initiation (Scenario 2): In the base case analysis, we accounted for the likelihood of initiating treatment after five days of symptom onset for which we assumed zero treatment efficacy. To assess the impact of treatment initiation on our results, we modeled a scenario that assumed a 100% probability of early treatment initiation in all three study countries.
3. Inclusion of productivity losses (Scenario 3): A broader perspective was adopted to account for productivity losses in the acute phase of COVID-19 disease and due to premature death. Time lost from work was valued using each study country’s GDP per capita^34^ and adjusted for unemployment rates^35^ over a period equivalent to the duration of illness for surviving patients and up to retirement age ^36, 37^ for patients who die (Table 3). Productivity loses due to COVID-19 death were discounted using a 5% discount rate ^29^.
4. Post-acute impact of COVID-19 (Scenario 4): Patients with severe COVID-19 are likely to be readmitted to hospital following the initial acute phase of the disease ^38^. To account for this, we extended the model to account for readmission one year following the acute disease phase (Figure S2). We assumed one hospital readmission during this time and obtained relevant data on readmission rates and length of hospitalization from existing literature ^38^. Cost of readmission was assumed to be the average costs of inpatient care for COVID-19 while disability weight for post-acute disease phase where based on GBS 2019 disability weights recommended by the European Burden of Disease Network Consensus COVID-19 models ^28^ (Table 3).
5. Inclusion of future unrelated health care costs (Scenario 5): To account for future unrelated health care costs, all surviving cases were allocated an annual cost over their remaining life expectancy valued at the health expenditure per capita for each country^34^. All future unrelated health care costs were discounted using a 5% discount rate ^29^.

**Table 2:** Scenario description.

|  |  |
| --- | --- |
| Scenario 1 | Assesses cost-effectiveness in COVID-19 vaccinated patients |
| Scenario 2 | Assumes 100% probability of treatment initiation within 5 days of symptom onset |
| Scenario 3 | Accounts for productivity losses due to COVID-19 illness and premature death |
| Scenario 4 | Accounts for readmissions in the first year following the acute disease phase |
| Scenario 5 | Accounts for future unrelated healthcare costs |

**Table 3:** Model Parameters (Scenario analyses)

| Parameters | Ghana | Rwanda | Zambia |
| --- | --- | --- | --- |
| **Vaccine effectiveness** | 0.972 | 0.972 | 0.972 |
| **Vaccination coverage** | 0.35 | 0.68 | 0.12 |
| **Vaccine weighted Inpatient care** |  |  |  |
| Inpatient care (all adult population) | 0.018 | 0.0045 | 0.0177 |
| Inpatient care (adults $\geq 65$ years old) | 0.162 | 0.0407 | 0.159 |
| Inpatient care (adults with other risk factors ) | 0.0541 | 0.0136 | 0.0530 |
| **Treatment effect in vaccinated individuals (Paxlovid)** |  |  |  |
| Relative risk, hospitalisation | 0.58 | 0.58 | 0.58 |
| **Productivity loss: due to acute illness (US\$)** | | | |
| Outpatient care only | 28.80 | 8.36 | 12.65 |
| Mild/moderate, survived | 17.40 | 5.05 | 7.64 |
| Mild/moderate, died | 29.82 | 8.66 | 13.10 |
| Severe, survived | 29.58 | 8.59 | 12.99 |
| Severe, died | 17.57 | 5.10 | 7.72 |
| Critical, survived | 29.58 | 8.59 | 12.99 |
| Critical, died | 17.57 | 5.10 | 7.72 |
| **Productivity loss: due to premature death (US\$)** | | | |
| All adult population | 24 774.47 | 10 462.95 | 10 883.96 |
| Adults ≥ 65 years old | 0 | 0 | 0 |
| Adults with comorbidities | 24 774.47 | 10 462.95 | 10 883.96 |
| GDP per capita (US\$) | 2 363.30 | 822.30 | 1 137.30 |
| Unemployment rate | 0.047 | 0.205 | 0.13 |
| Compulsory retirement age (years) | 60 | 65 | 60 |
| **Model extension** |  |  |  |
| Proportion readmitted | 0.069 | 0.069 | 0.069 |
| Number of readmissions | 1 | 1 | 1 |
| Readmission length of hospital stay | 12 | 12 | 12 |
| Mortality risk following survival with long-term sequelae | 0.04 | 0.04 | 0.04 |
| Disability weight: Post-acute of COVID-19 | 0.22 | 0.22 | 0.22 |
| Disability duration (years) | 1 | 1 | 1 |
| Cost of readmissions (US\$) | 1 171.75 | 1 066.11 | 1 181.00 |
| **Future unrelated health care costs (US\$)** | | | |
| All adult population | 3 022.74 | 1 969.91 | 2 899.33 |
| Adults ≥ 65 years old | 1 492.38 | 970.22 | 1 475.18 |
| Adults with comorbidities | 3 022.74 | 1 969.91 | 2 899.33 |

## Results

### Base case

Table 4 summarizes the results of the base case analysis. The results varied across the three countries and target populations. – In the elderly population (Table 4, Panel B), Molnupiravir and Paxlovid were both less costly (indicated as negative incremental costs) and more effective (indicated as positive DALYs averted)) than the comparator (i.e., Molnupiravir and Paxlovid dominated usual care) in Rwanda and Zambia. In Ghana, Paxlovid dominated usual care (incremental costs =-US$20.75 and DALYs averted= 0.007, Table 4 Panel B) while Molnupiravir was both more costly (incremental cost= US$ 2.12, Table 4, Panel B) and more effective (DALYs averted = 0.002, Table 4, Panel B) resulting in an ICER of US$ 1023.58 per DALY averted (Table 4, Panel B).

**Table 4:**
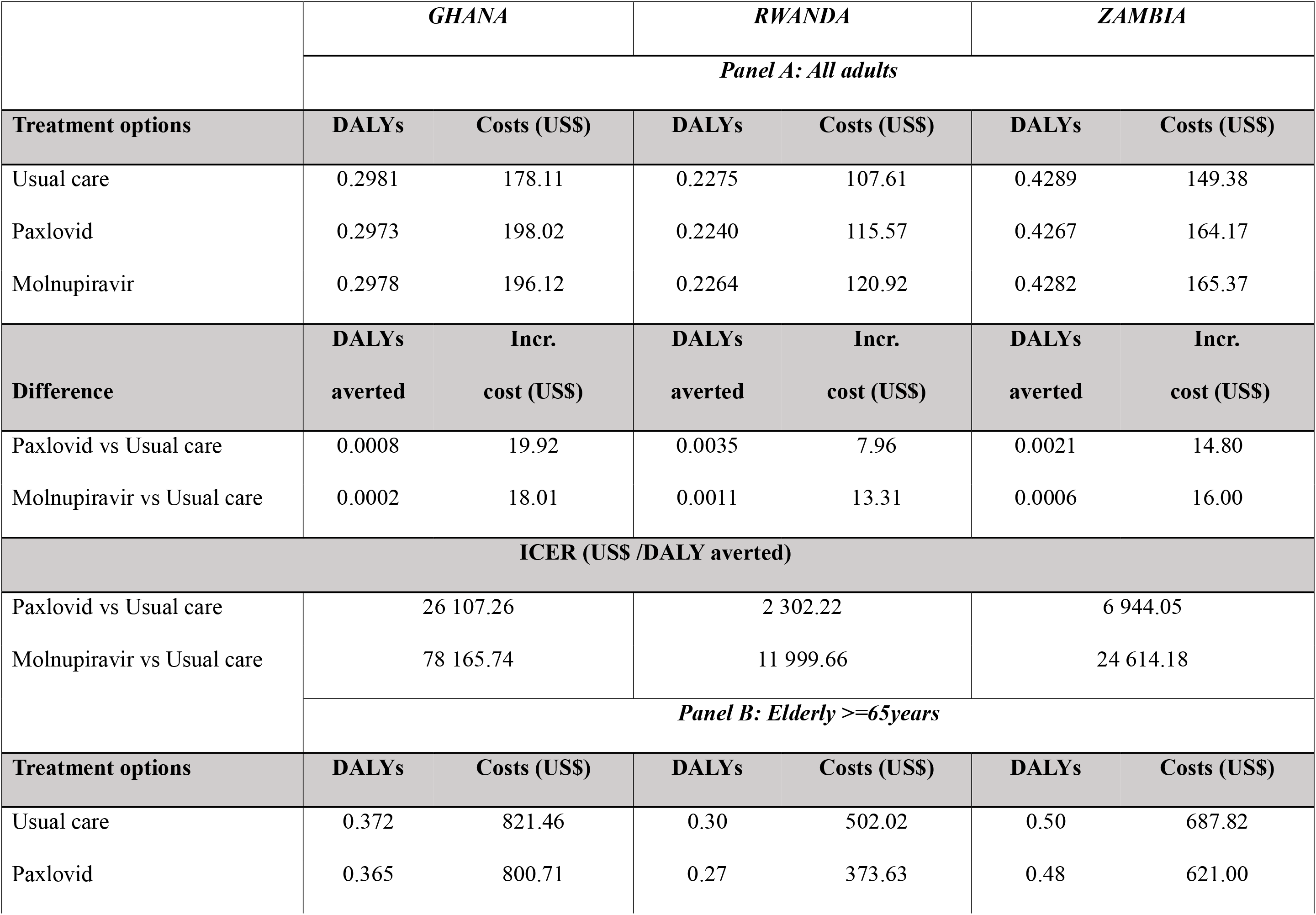
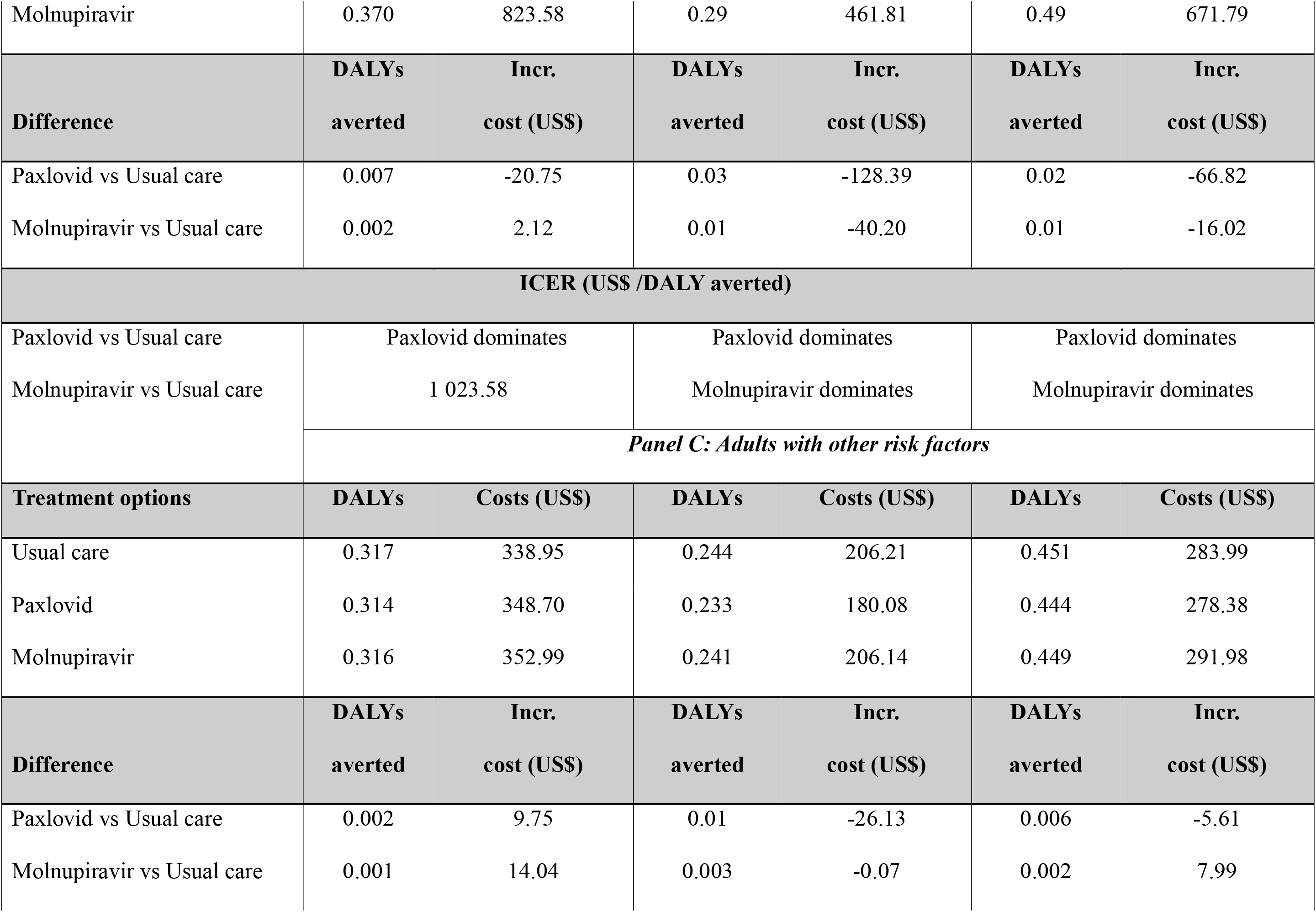
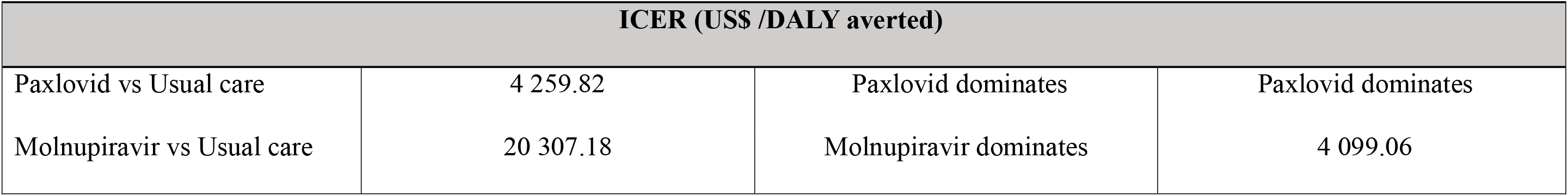
Incremental outcomes and incremental cost-effectiveness ratios (Base case)

|  | GHANA |  | RWANDA |  | ZAMBIA |  |
| --- | --- | --- | --- | --- | --- | --- |
|  | Panel A: All adults |  |  |  |  |  |
| Treatment options | DALYs | Costs (US\$) | DALYs | Costs (US\$) | DALYs | Costs (US\$) |
| Usual care | 0.2981 | 178.11 | 0.2275 | 107.61 | 0.4289 | 149.38 |
| Paxlovid | 0.2973 | 198.02 | 0.2240 | 115.57 | 0.4267 | 164.17 |
| Molnupiravir | 0.2978 | 196.12 | 0.2264 | 120.92 | 0.4282 | 165.37 |
| Difference | DALYs | Incr. | DALYs | Incr. | DALYs | Incr. |
| | averted | cost (US\$) | averted | cost (US\$) | averted | cost (US\$) |
| Paxlovid vs Usual care | 0.0008 | 19.92 | 0.0035 | 7.96 | 0.0021 | 14.80 |
| Molnupiravir vs Usual care | 0.0002 | 18.01 | 0.0011 | 13.31 | 0.0006 | 16.00 |
| ICER (US\$ /DALY averted) | | | | | | |
| Paxlovid vs Usual care | 26 107.26 |  | 2 302.22 |  | 6 944.05 |  |
| Molnupiravir vs Usual care | 78 165.74 |  | 11 999.66 |  | 24 614.18 |  |
|  | Panel B: Elderly >=65years |  |  |  |  |  |
| | DALYs | Costs (US\$) | DALYs | Costs (US\$) | DALYs | Costs (US\$) |
| Usual care | 0.372 | 821.46 | 0.30 | 502.02 | 0.50 | 687.82 |
| Paxlovid | 0.365 | 800.71 | 0.27 | 373.63 | 0.48 | 621.00 |
| Molnupiravir | 0.370 | 823.58 | 0.29 | 461.81 | 0.49 | 671.79 |
|  | DALYs | Incr. | DALYs | Incr. | DALYs | Incr. |
| Difference | averted | cost (US\$) | averted | cost (US\$) | averted | cost (US\$) |
| Paxlovid vs Usual care | 0.007 | -20.75 | 0.03 | -128.39 | 0.02 | -66.82 |
| Molnupiravir vs Usual care | 0.002 | 2.12 | 0.01 | -40.20 | 0.01 | -16.02 |
| ICER (US\$ /DALY averted) | | | | | | |
| Paxlovid vs Usual care | Paxlovid dominates |  | Paxlovid dominates |  | Paxlovid dominates |  |
| Molnupiravir vs Usual care | 1 023.58 |  | Molnupiravir dominates |  | Molnupiravir dominates |  |
|  | Panel C: Adults with other risk factors |  |  |  |  |  |
| Treatment options | DALYs | Costs (US\$) | DALYs | Costs (US\$) | DALYs | Costs (US\$) |
| Usual care | 0.317 | 338.95 | 0.244 | 206.21 | 0.451 | 283.99 |
| Paxlovid | 0.314 | 348.70 | 0.233 | 180.08 | 0.444 | 278.38 |
| Molnupiravir | 0.316 | 352.99 | 0.241 | 206.14 | 0.449 | 291.98 |
|  | DALYs | Incr. | DALYs | Incr. | DALYs | Incr. |
| Difference | averted | cost (US\$) | averted | cost (US\$) | averted | cost (US\$) |
| Paxlovid vs Usual care | 0.002 | 9.75 | 0.01 | -26.13 | 0.006 | -5.61 |
| Molnupiravir vs Usual care | 0.001 | 14.04 | 0.003 | -0.07 | 0.002 | 7.99 |

| ICER (US\$ /DALY averted) | | | |
| --- | --- | --- | --- |
| Paxlovid vs Usual care | 4 259.82 | Paxlovid dominates | Paxlovid dominates |
| Molnupiravir vs Usual care | 20 307.18 | Molnupiravir dominates | 4 099.06 |

In adults with other risk factors (Table 4, Panel C), Paxlovid dominated in Rwanda and Zambia, and was both more costly (incremental cost= US$9.75) and more effective (DALYs averted = 0.002) in Ghana, resulting in an estimated ICER of US$ 4259.82 per DALY averted (Table 4, Panel C); Molnupiravir dominated only in Rwanda, and in Ghana and Zambia, ICERs were estimated at US$ 20 307.18 and US$4 099.06 per DALY averted, respectively (Table 4, Panel C).

In the all-adult patient target population, across all three countries Paxlovid and Molnupiravir were both more costly and more effective resulting in ICERs ranging from US$ 2302.22 per DALY averted in Rwanda to US$ 26 107.26 in Ghana for Paxlovid and from US$ 11 999.66 per DALY averted in Rwanda to US$ 78 165.74 in Ghana for Molnupiravir (Table 4, Panel A).

Figure 2 presents INMBs of Paxlovid and Molnupiravir compared to usual care estimated at the cost-effectiveness threshold assumed for each country (Table 1). Except for the all-adult target population in Ghana, Paxlovid is observed to have a higher INMB compared to Molnupiravir across all target populations and study countries (Figure 2).

**Figure 2:**
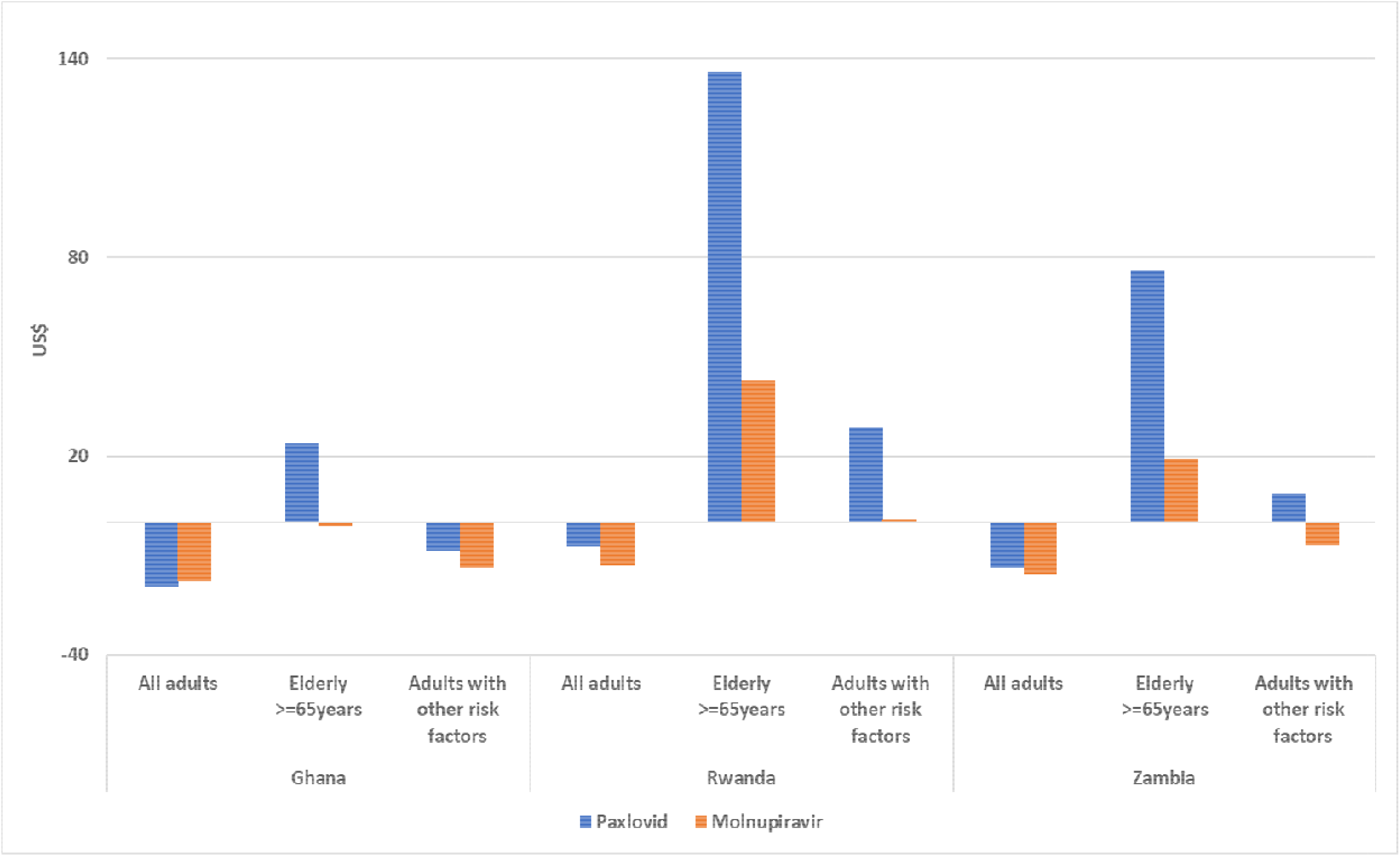
Incremental Net Monetary Benefits*,Paxlovid and Molnupiravir compared to Usual care.

### Sensitivity analysis

The results of the one-way deterministic sensitivity analyses, estimated using INMBs, are shown in Figure S3. Across all study countries, three parameters had the largest impact on the base results for Paxlovid –COAV treatment costs, the likelihood of treatment initiation within five days of symptom onset, and hospitalization rate modifiers affecting hospitalization rates for the elderly population and adults with other risk factors (Figure S3). For example, in Ghana, at a lower hospitalisation rate multiplier in the elderly population (1.8 vs 9 in the base case, Table S2), the INMB for Paxlovid became negative at a cost-effectiveness threshold of US$430 per DALY averted while at a higher hospitalisation rate multiplier (16.2 vs 9 in the base case, Table S2), the INMB for Paxlovid became positive in adults with other risk factors. At lower Paxlovid treatment cost (US$5 vs US$ 25 in the base case, Table S2), the INMB for Paxlovid in Ghana became positive in the all-adult target population and in adults with other high-risk factors for disease progression. In Rwanda and Zambia, Paxlovid treatment cost, hospitalization rate modifiers and the likelihood of early treatment initiation similarly have the largest impact on the base case INMBs.

Across the three study countries and target populations, uncertainty in model parameters followed a similar pattern for Molnupiravir where COAV treatment cost, likelihood of early treatment initiation, hospitalisation rate modifier and in addition, Molnupiravir treatment effect had the largest impact on the base case results for Molnupiravir (Figure S3).

The results of the probabilistic sensitivity analysis are summarized in cost-effectiveness acceptability curves (Figure 3-5). Compared to other target populations, the proportion of simulations that were cost-effective was highest in the elderly population for both Molnupiravir and Paxlovid (Figures 4). Conversely, in the all-adult population, the proportion of simulations that were cost-effective for Paxlovid and Molnupiravir were very low across all study countries-approximately 0% for Molnupiravir in all study counties and 0%, <10% and 0% for Paxlovid in Ghana, Rwanda, and Zambia, respectively, at cost-effectiveness thresholds assumed for each country (Figure 3). In patients with other risk factors, the PSA results were also mixed – across study countries, the proportion of simulations falling below the cost-effectiveness threshold assumed was 0%, <45% and <10% for Molnupiravir in Ghana, Rwanda, and Zambia, respectively and <10%, approximately100% and <80% for Paxlovid in Ghana, Rwanda and Zambia, respectively (Figure 5).

**Figure 3:**
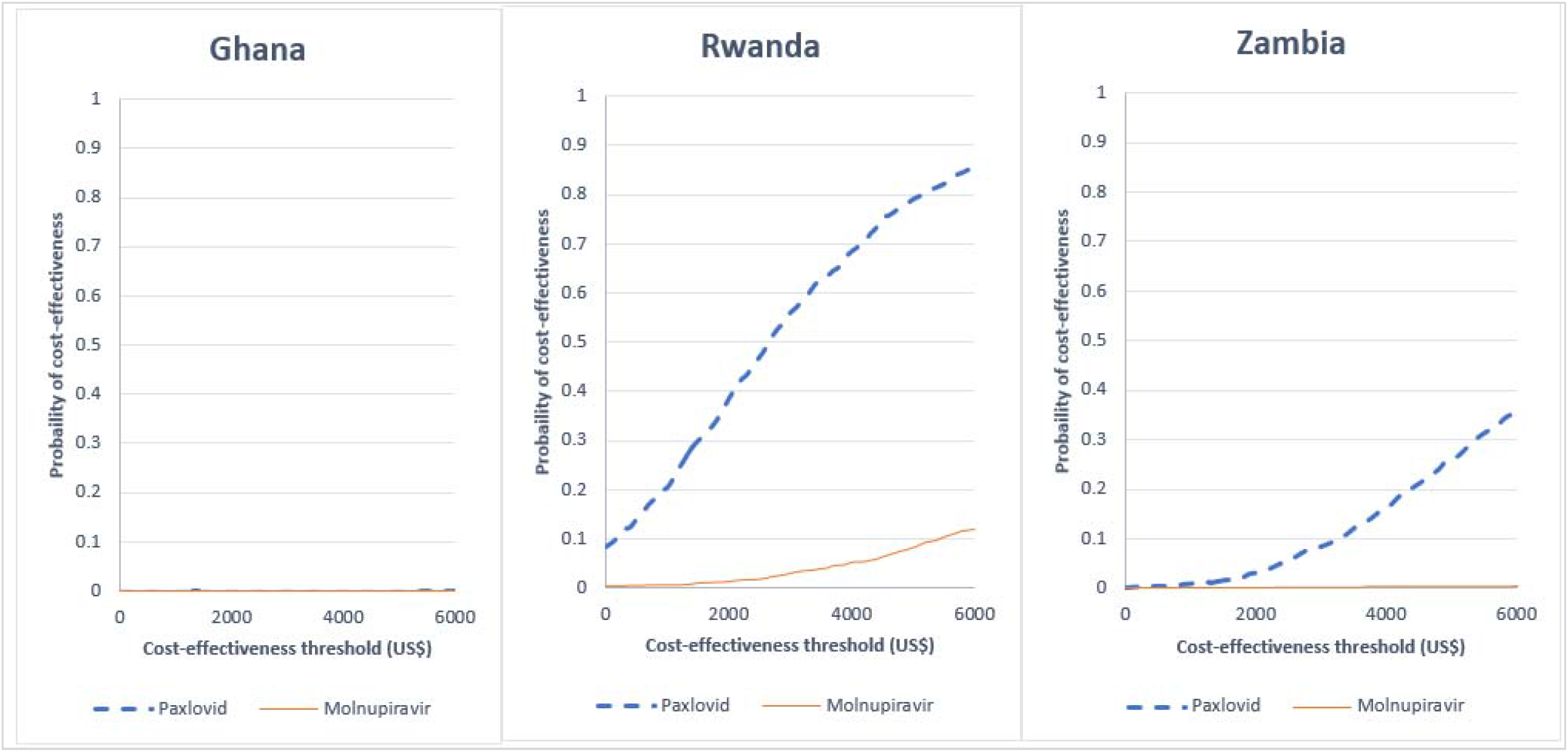
Cost-effectiveness acceptability curves (general adult population)

**Figure 4:**
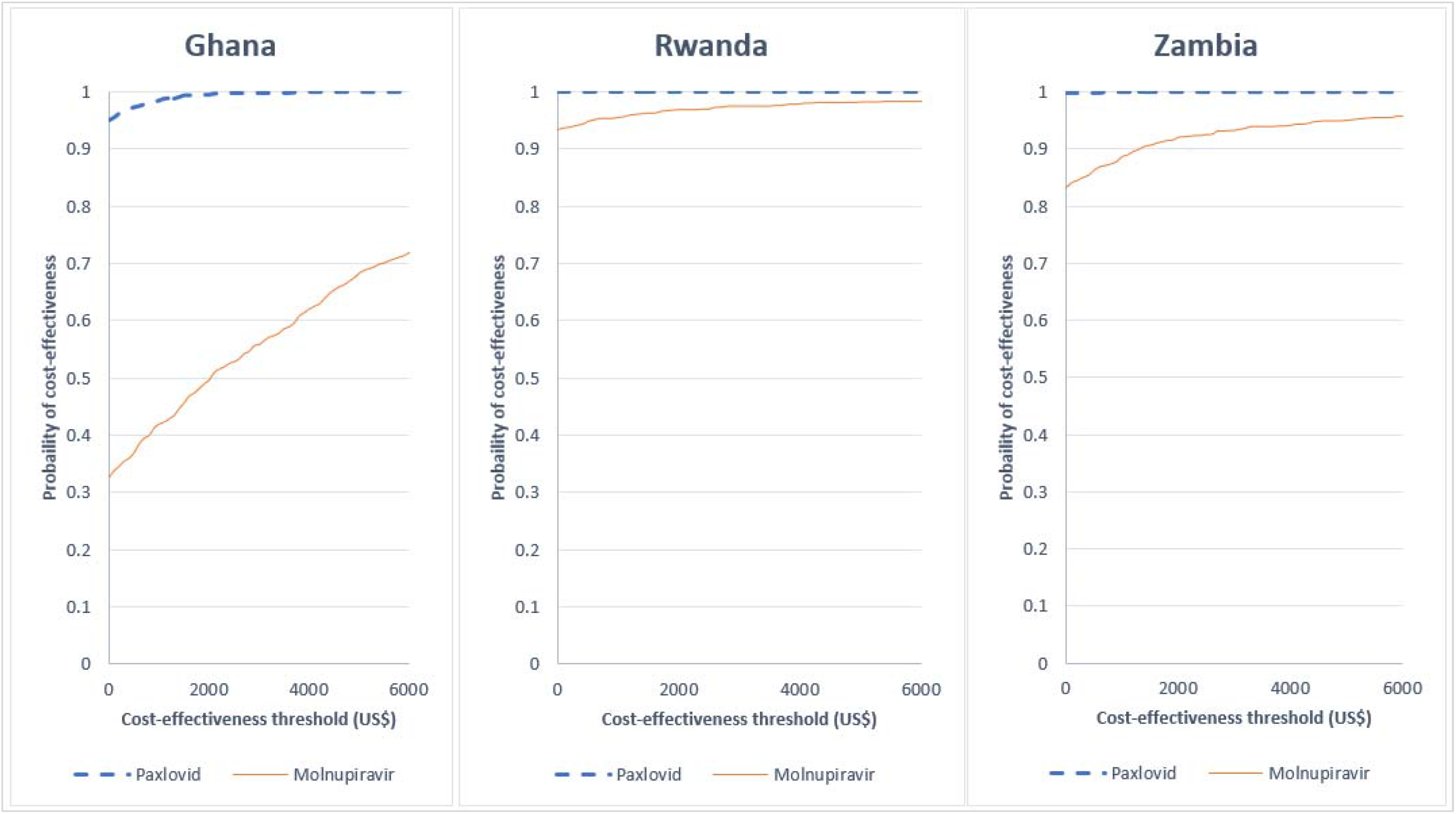
Cost-effectiveness acceptability curves (elderly population)

**Figure 5:**
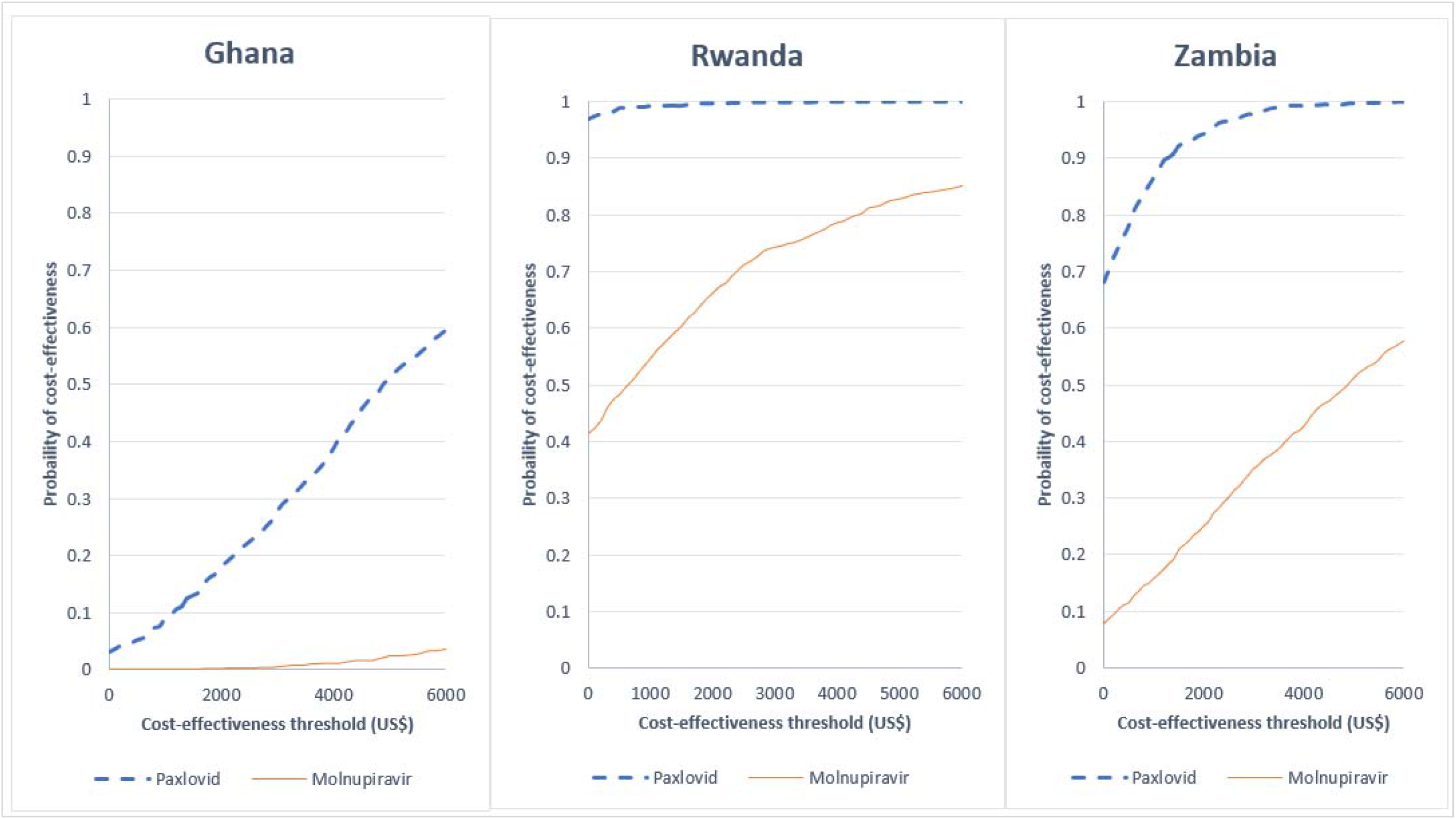
Cost-effectiveness acceptability curves (other risk groups)

### Scenario Analysis

The results of the scenario analysis are displayed in Table S3. Across all three countries and target populations modelled, the results for Paxlovid and Molnupiravir are largely unaffected by the inclusion of productivity losses, model extension to account for readmission and the inclusion of future unrelated health costs (Table S3). However, the results differed substantially from the base-case analysis for scenarios 1 (COVID-19 vaccine effectiveness) and 2 (100% probability of early treatment initiation). When COVID-19 vaccine effectiveness is accounted for in the model (scenario 1), the ICERs for Paxlovid for all study countries increased across all target populations, with the INMB remaining positive only for elderly patients in Rwanda and Zambia. When a 100% probability of early treatment is assumed in scenario 2, Paxlovid and Molnupiravir dominated usual care in all three study countries and in all target populations modeled except for Molnupiravir in the all-adult target population in Rwanda.

## Discussion

In this study, we modeled the effect of Paxlovid and Molnupiravir compared to usual care in three target populations and assessed scenarios and parameters that may affect the cost-effectiveness of both COAVs in the African setting.

Our results suggest that both Molnupiravir and Paxlovid could be cost-effective when unvaccinated, patients at high risk of disease progression are targeted. Our findings are significantly affected by key model parameters that varied across the three study countries modeled. For example, in Ghana and Zambia where the likelihood of early treatment initiation was assumed to be 7% and 18% respectively, Molnupiravir was not cost-effective in patients with other high-risk factors but dominated usual care in Rwanda where a higher early treatment initiation likelihood (41%) was assumed. The results are also sensitive to baseline hospitalization rates modeled. In elderly patients for whom we assumed the highest risk of disease progression, both Paxlovid and Molnupiravir are cost-saving across all study countries regardless of likelihood of early treatment initiation, except for in Ghana, where Molnupiravir was not cost-effective at the cost-effective threshold assumed. Neither Paxlovid nor Molnupiravir are cost-effective in the unvaccinated all-adult target population across all study countries. This is likely due to the lower risk of hospitalization modeled in this target population. However, when a 100% likelihood of early treatment initiation was assumed, both Molnupiravir and Paxlovid was cost-saving in the unvaccinated all-adult target population for all three study countries except in Rwanda, which had the lowest baseline hospitalization rate modeled. However, the finding in the all-adult population should be interpreted with caution given that evidence of treatment efficacy for both Molnupiravir and Paxlovid is restricted to populations at high risk of disease progression^6, 7^.

In a scenario that assessed cost-effectiveness in vaccinated patients, our finding suggests that Paxlovid could be cost-effective but only in vaccinated elderly patients, particularly in settings with higher likelihoods of treatment initiation within 5 days of symptom onset as seen in Rwanda and Zambia. The reduced effectiveness of Paxlovid in vaccinated patients^33^, combined with baseline hospitalization rates modeled may explain why Paxlovid was not cost-effective in adults with other high-risk factors for disease progression, contrary to results observed in unvaccinated individuals. However, the cost-effectiveness of Paxlovid in elderly population is based on effectiveness estimates drawn from an observational study^33^, which may be subject to higher degree of bias due to non-randomization of patients. Therefore, these findings should be reassessed as new evidence from RCTs emerges.

Paxlovid was observed to consistently have a higher INMB than the INMB for Molnupiravir. This suggests that the additional value generated per additional cost is higher for Paxlovid. However, this conclusion has an important caveat. There are no head-to-head RCTs assessing the efficacy of Molnupiravir vs Paxlovid under consistent trial conditions. Differences between the Molnupiravir and Paxlovid RCT settings and study participants’ characteristics, such as differences in the standard of care, predominant COVID-19 variant and previous exposure to COVID-19^1^ limits the usefulness of a direct comparison of the effectiveness from the individual RCTs and thus, the cost-effectiveness of Paxlovid vs Molnupiravir.

Our findings are based on the assumption that both Paxlovid and Molnupiravir could be accessed at lower prices. However, as highlighted in the DSA, COAV treatment costs could impact on the findings reported here such that at treatment costs similar to high income countries, Paxlovid (at approximately $600 per treatment course) and Molnupiravir (at approximately $700 per treatment course) will not be cost-effective for any target population in our study setting.

From the probabilistic sensitivity analysis, our base-case findings are robust to joint uncertainty in model parameters but to a lesser extent for Molnupiravir compared to Paxlovid – Paxlovid had a higher probability of being cost-effective compared to the probability of Molnupiravir being cost-effective across all study countries.

Notwithstanding contextual differences between settings, our findings are consistent with findings from other settings where Paxlovid and Molnupiravir each compared to usual care, have been shown to be cost-effective in high-risk populations with mild/moderate COVID-19. Despite comparatively lower hospitalization rates seen in Africa, Paxlovid could be cost-effective in limited use cases, if offered at lower prices. Given the lower vaccination coverage observed in Africa, these treatment regimens could be beneficial for unvaccinated sub-populations at high risk of disease progression.

This study has some limitations. First, our model was designed to assess the cost-effectiveness of each COAV compared to usual care. As with previous studies ^10, 12, 39^, a head-to-head comparison of Paxlovid and Molnupiravir was not conducted due to differences in RCT design features and populations. Second, due to a dearth of context-specific data, the study relied on proxies and plausible assumptions for key model parameters – hospitalization rates were largely drawn from studies in the first year of the pandemic. However, in the current era of the milder Omicron variant and high natural immunity, COVID-19 hospitalization rates have substantially decreased across all settings. Although we accounted for this by adjusting hospitalization rates downward, we cannot rule out that our base case hospitalization rates remain too high. Context-specific costs of the clinical management of COVID-19 were not available for Rwanda and Zambia. Therefore, we relied on costs estimates from a large multi-country extrapolation study ^26^. Although we adjusted these cost estimates to be more comparable to clinical management cost from single-country studies, this may have resulted in a biased estimate of the disease management cost in Rwanda and Zambia. Unlike previous cost-effectiveness analyses of COAVs, this study explicitly modelled the implication of early treatment initiation on the cost-effectiveness of COAVs. However, we used the total COVID-19 test performed per 1000 population as a proxy for the likelihood of early treatment initiation. While testing rates are useful indicators of the stringency of COVID-19 control measures within countries, without knowledge of the timing of testing, it is unclear if test rates themselves suitably capture the probability of early diagnosis and treatment initiation.

## Conclusion

Paxlovid has the potential to be a cost-effective treatment option in African countries similar to Ghana, Rwanda or Zambia for the limited use case of unvaccinated patients at high risk of severe disease progression. More evidence is needed to determine cost-effectiveness for patients that are unvaccinated but have previously been infected with COVID-19 and may have developed some immune protection. While this evidence is based on three countries, the diversity of key parameters across the three study countries allowed for explicit modeling of nuances relevant to the African setting. Therefore, this study provides useful insights to policymakers on a range of context-specific factors that should be considered when making funding decisions on COAVs based on findings from cost-effectiveness analyses.

## Data Availability

All data produced in the present work are contained in the manuscript

## Footnotes

1 For example, participants with previous exposure to COVID-19 were excluded from the RCT for Paxlovid but were included in the RCT for Molnupiravir.

