## Supplemental materials for "A cost-effectiveness analysis of Molnupiravir and Paxlovid in three African countries"

Supplementary Material

Table S1: Test performed per 1000 population, 2022 (month, REF)

| **200-1000** | **100-200** | **0-100** |
| --- | --- | --- |
| Gabon | Zimbabwe | Kenya |
| Mauritius | **Zambia** | Uganda |
| South Africa | Burundi | Togo |
| Namibia | Cameroon | **Ghana** |
| Mauritania |  | Cote d'Ivoire |
| Equatorial Guinea |  | Guinea |
| **Rwanda** |  | Guinea-Bissau |
| Eswatini |  | Mali |
| Lesotho |  | Nigeria |
|  |  | South Sudan |
|  |  | Ethiopia |
|  |  | Somalia |
|  |  | Madagascar |
|  |  | Malawi |
|  |  | Mozambique |
|  |  | Central Africa Republic |
|  |  | Chad |

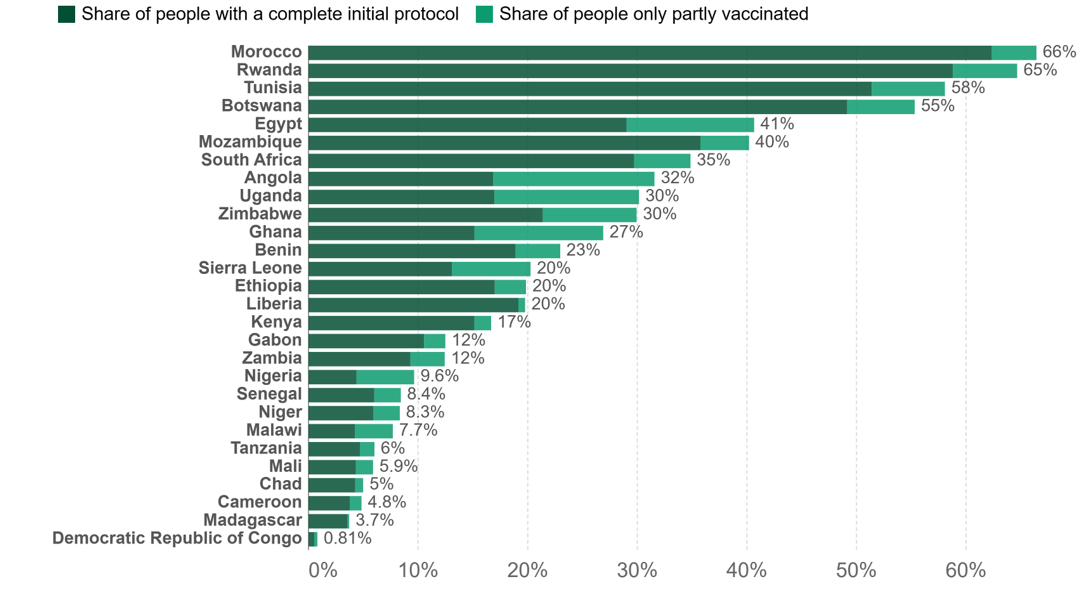

Figure S1: Share of population vaccinated against COVID-19 (March 2022), REF

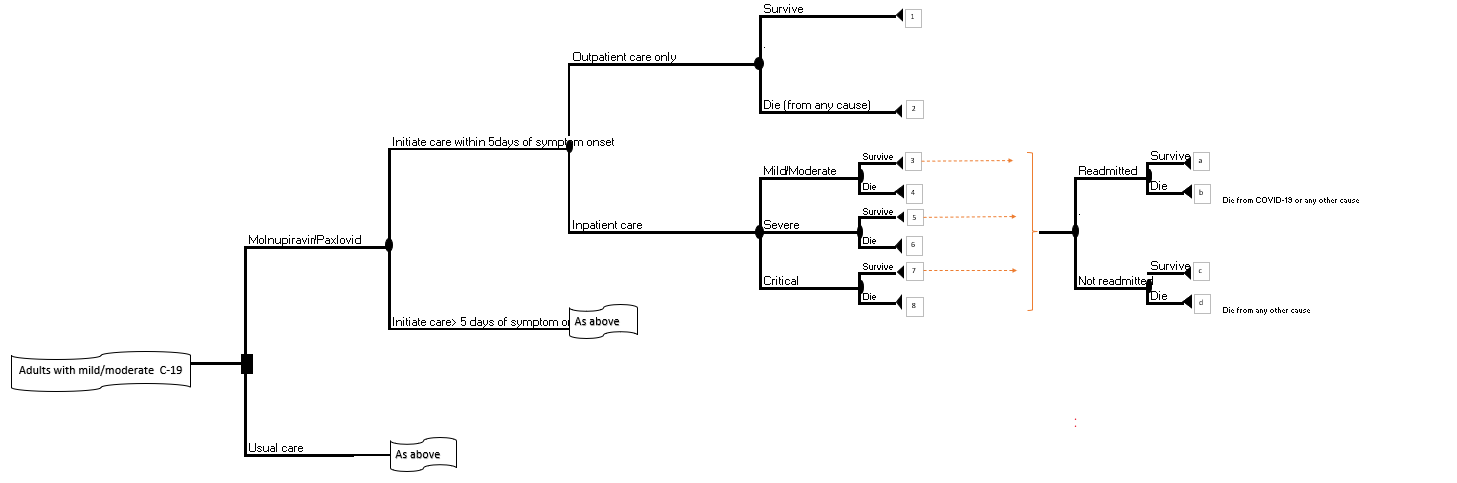

Figure S2: Model structure depicting hospital readmission post-acute COVID-19 phase.

**Table S2**: One-Way Deterministic Sensitivity Analysis Parameters

|  | **Parameters** | **Ghana** |  |  | **Rwanda** |  |  | **Zambia** |  |  |
| --- | --- | --- | --- | --- | --- | --- | --- | --- | --- | --- |
|  |  | Base  case | Low  value | High value | Base  case | Low value | High value | Base  case | Low value | High value |
|  | Discount rate | 0.05 | 0 | 0.1 | 0.05 | 0 | 0.1 | 0.05 | 0 | 0.1 |
| 1 | **Treatment effect (Paxlovid)** |  |  |  |  |  |  |  |  |  |
|  | RR, hospitalisation | 0.15 | 0.08 | 0.29 | 0.15 | 0.08 | 0.29 | 0.15 | 0.08 | 0.29 |
|  | RR, mortality in hospitalised patients | 0.12 | 0.04 | 0.36 | 0.12 | 0.04 | 0.36 | 0.12 | 0.04 | 0.36 |
| 2 | **Treatment effect (Molnupiravir)** |  |  |  |  |  |  |  |  |  |
|  | RR, hospitalisation | 0.67 | 0.49 | 0.92 | 0.67 | 0.49 | 0.92 | 0.67 | 0.49 | 0.92 |
| 3 | **COAV treatment costs** |  |  |  |  |  |  |  |  |  |
|  | Paxlovid | 25 | 5 | 45 | 25 | 5 | 45 | 25 | 5 | 45 |
|  | Molnupiravir | 20 | 4 | 36 | 20 | 4 | 36 | 20 | 4 | 36 |
| 4 | **Disease parameters** |  |  |  |  |  |  |  |  |  |
|  | %Receiving care within 5 days | 0.075 | 0.046 | 0.104 | 0.411 | 0.250 | 0.572 | 0.180 | 0.110 | 0.251 |
|  | % Inpatient care | 0.082 | 0.050 | 0.114 | 0.040 | 0.024 | 0.056 | 0.060 | 0.036 | 0.084 |
|  | Hospitalisation modifier (Age) | 9 | 1.8 | 16.2 | 9 | 1.8 | 16.2 | 9 | 1.8 | 16.2 |
|  | Hospitalisation modifier (Comorbidities) | 3 | 0.6 | 5.4 | 3 | 0.6 | 5.4 | 3 | 0.6 | 5.4 |
| 5 | **Mortality rates** |  |  |  |  |  |  |  |  |  |
|  | Outpatient care only | 0.039 | 0.024 | 0.054 | 0.030 | 0.018 | 0.042 | 0.055 | 0.033 | 0.076 |
|  | Mild/moderate | 0.039 | 0.024 | 0.054 | 0.030 | 0.018 | 0.042 | 0.055 | 0.033 | 0.076 |
|  | Severe | 0.171 | 0.104 | 0.239 | 0.207 | 0.126 | 0.288 | 0.189 | 0.115 | 0.264 |
|  | Critical | 0.257 | 0.156 | 0.358 | 0.207 | 0.126 | 0.288 | 0.338 | 0.206 | 0.471 |
| 6 | **Cost of Illness (US$), per patient per day** |  |  |  |  |  |  |  |  |  |
|  | Outpatient care only | $ 20.93 | $ 12.73 | $ 29.14 | $ 12.49 | $ 7.60 | $ 17.39 | $ 17.59 | $ 10.69 | $ 24.48 |
|  | Mild/moderate | $ 581.19 | $ 353.36 | $ 809.01 | $ 502.41 | $ 305.46 | $ 699.35 | $ 583.45 | $ 354.74 | $ 812.16 |
|  | Severe | $ 1363.70 | $ 829.13 | $1898.28 | $ 1178.85 | $ 716.74 | $1640.97 | $1369.00 | $ 832.35 | $1905.65 |
|  | Critical | $ 1 570.36 | $ 954.78 | $2 185.94 | $ 1 517.06 | $ 922.37 | $2 111.75 | $1 590.55 | $ 967.06 | $2 214.05 |
| 7 | **Duration of care (Los)** |  |  |  |  |  |  |  |  |  |
|  | Outpatient care only | 5 | 3 | 6 | 5 | 3 | 6 | 5 | 3 | 6 |
|  | Mild/moderate, survived | 3 | 3 | 3 | 3 | 3 | 3 | 3 | 3 | 3 |
|  | Mild/moderate, died | 5 | 5 | 5 | 5 | 5 | 5 | 5 | 5 | 5 |
|  | Severe, survived | 5 | 4 | 5 | 5 | 4 | 5 | 5 | 4 | 5 |
|  | Severe, died | 3 | 2 | 3 | 3 | 2 | 3 | 3 | 2 | 3 |
|  | Critical, survived | 5 | 4 | 5 | 5 | 4 | 5 | 5 | 4 | 5 |
|  | Critical, died | 3 | 2 | 3 | 3 | 2 | 3 | 3 | 2 | 3 |
| 10 | **Disability weight** |  |  |  |  |  |  |  |  |  |
|  | Outpatient care only | 0.051 | 0.032 | 0.074 | 0.051 | 0.032 | 0.074 | 0.051 | 0.032 | 0.074 |
|  | Mild/moderate | 0.133 | 0.088 | 0.19 | 0.133 | 0.088 | 0.19 | 0.133 | 0.088 | 0.19 |
|  | Severe | 0.655 | 0.579 | 0.727 | 0.655 | 0.579 | 0.727 | 0.655 | 0.579 | 0.727 |
|  | Critical | 0.655 | 0.579 | 0.727 | 0.655 | 0.579 | 0.727 | 0.655 | 0.579 | 0.727 |

**Figure S3:** One-Way Deterministic Sensitivity Analysis (Base case)

1. **GHANA**

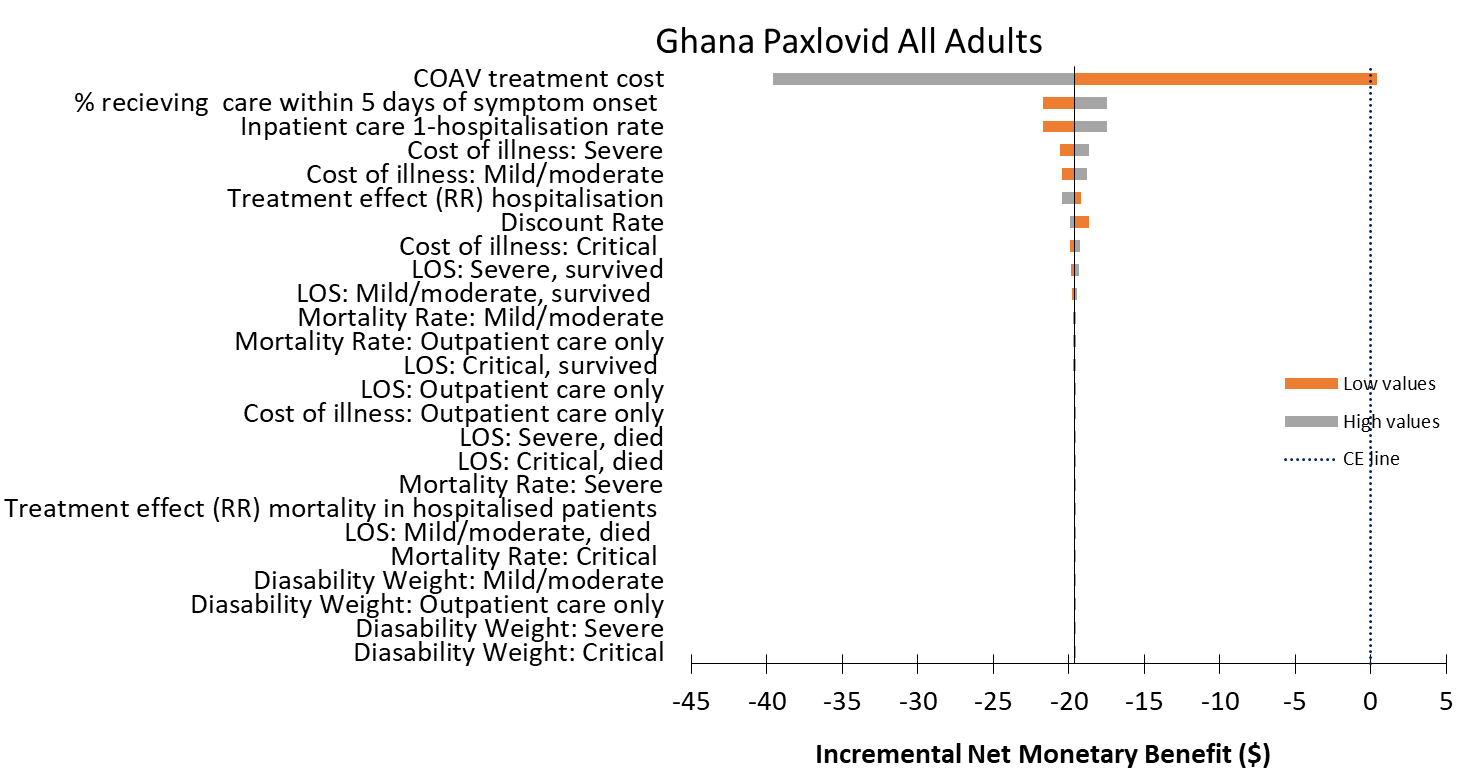

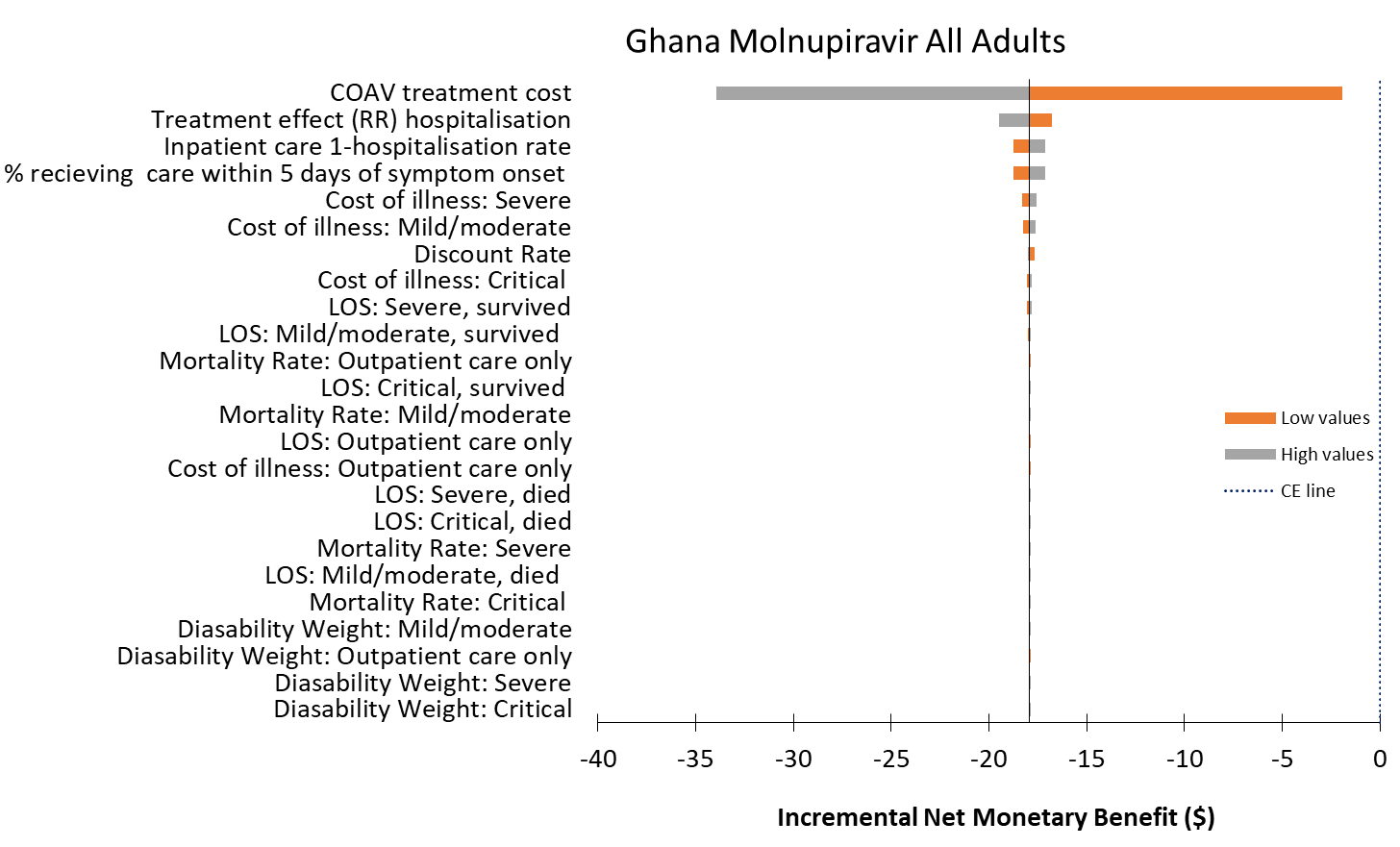

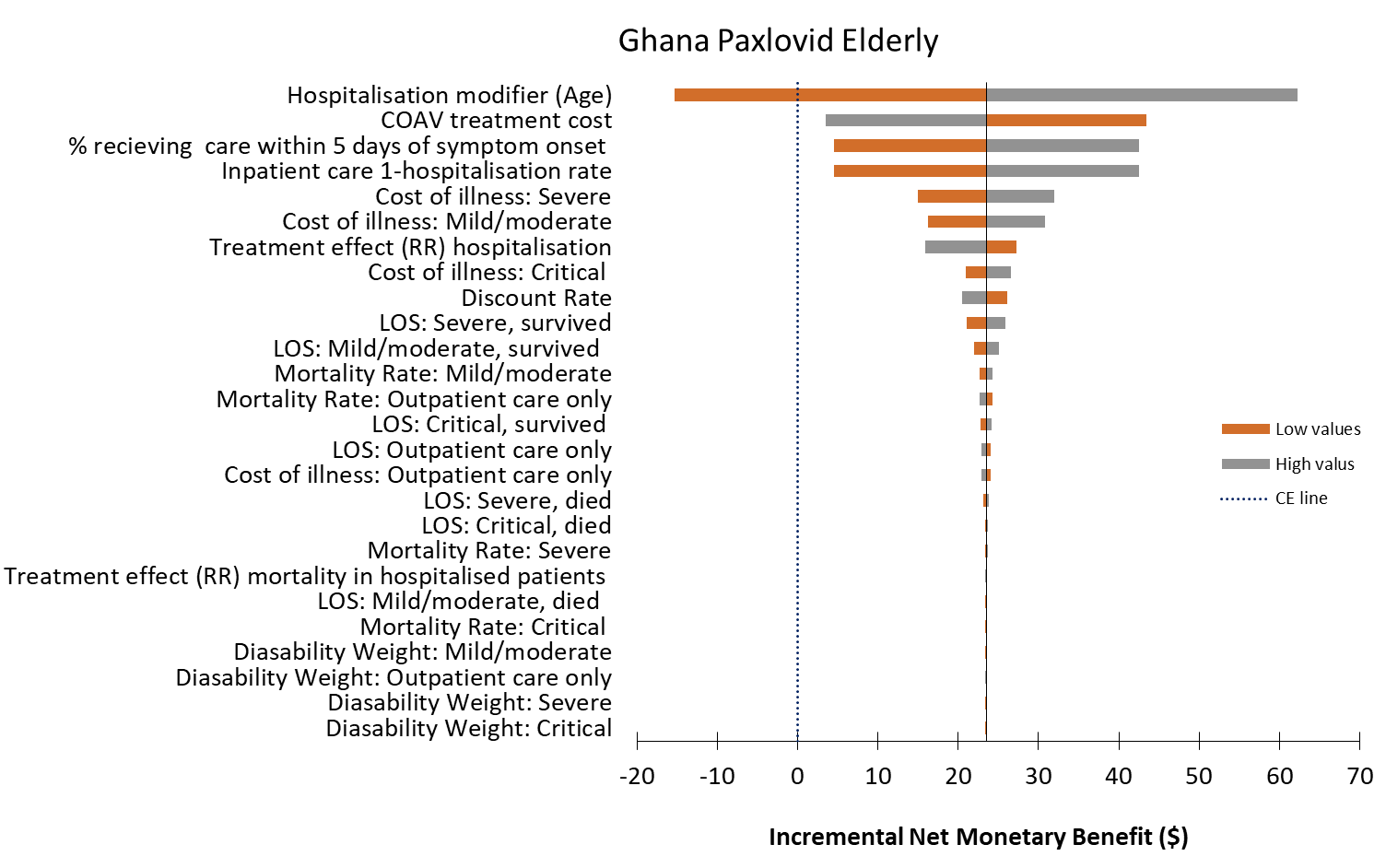

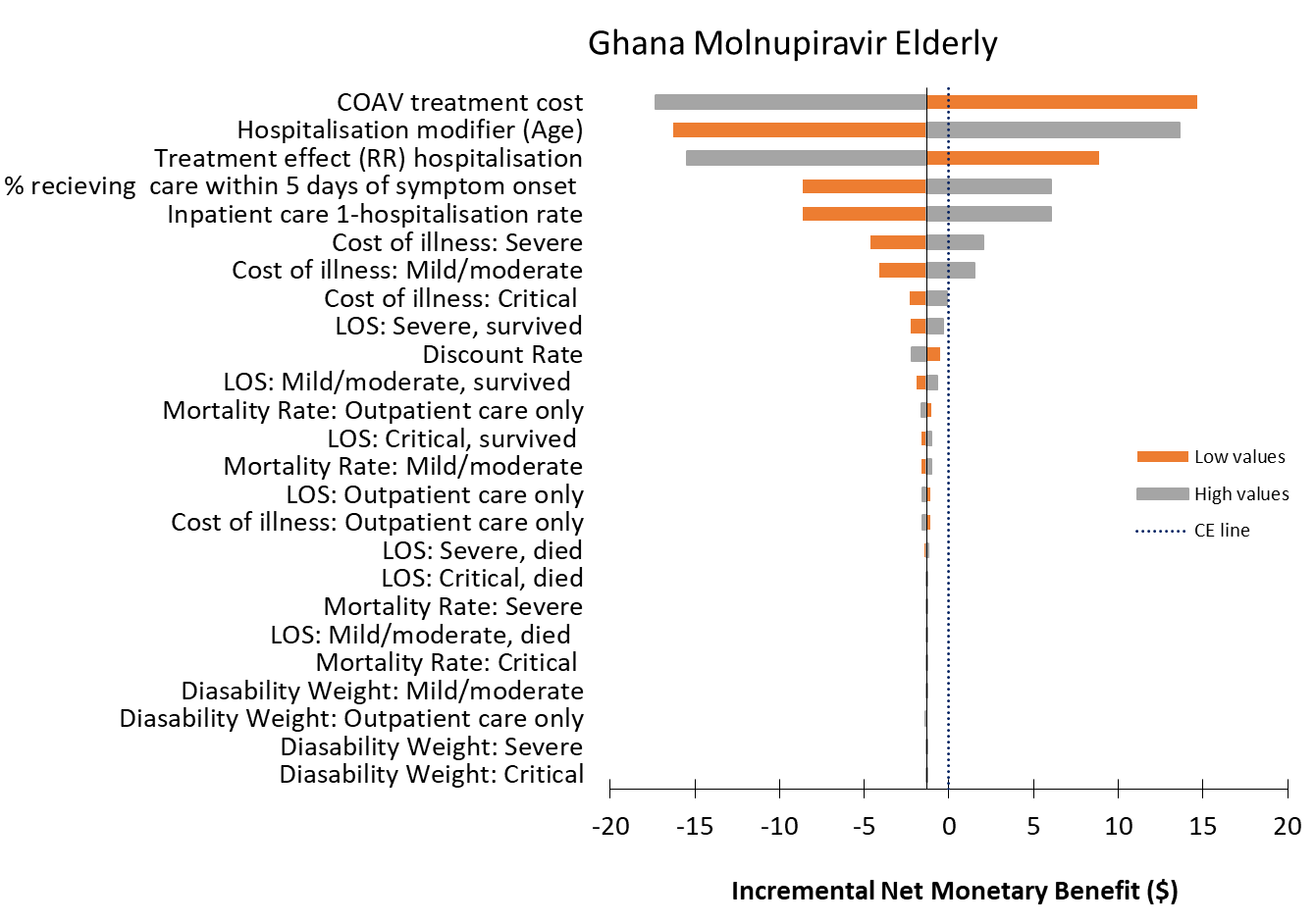

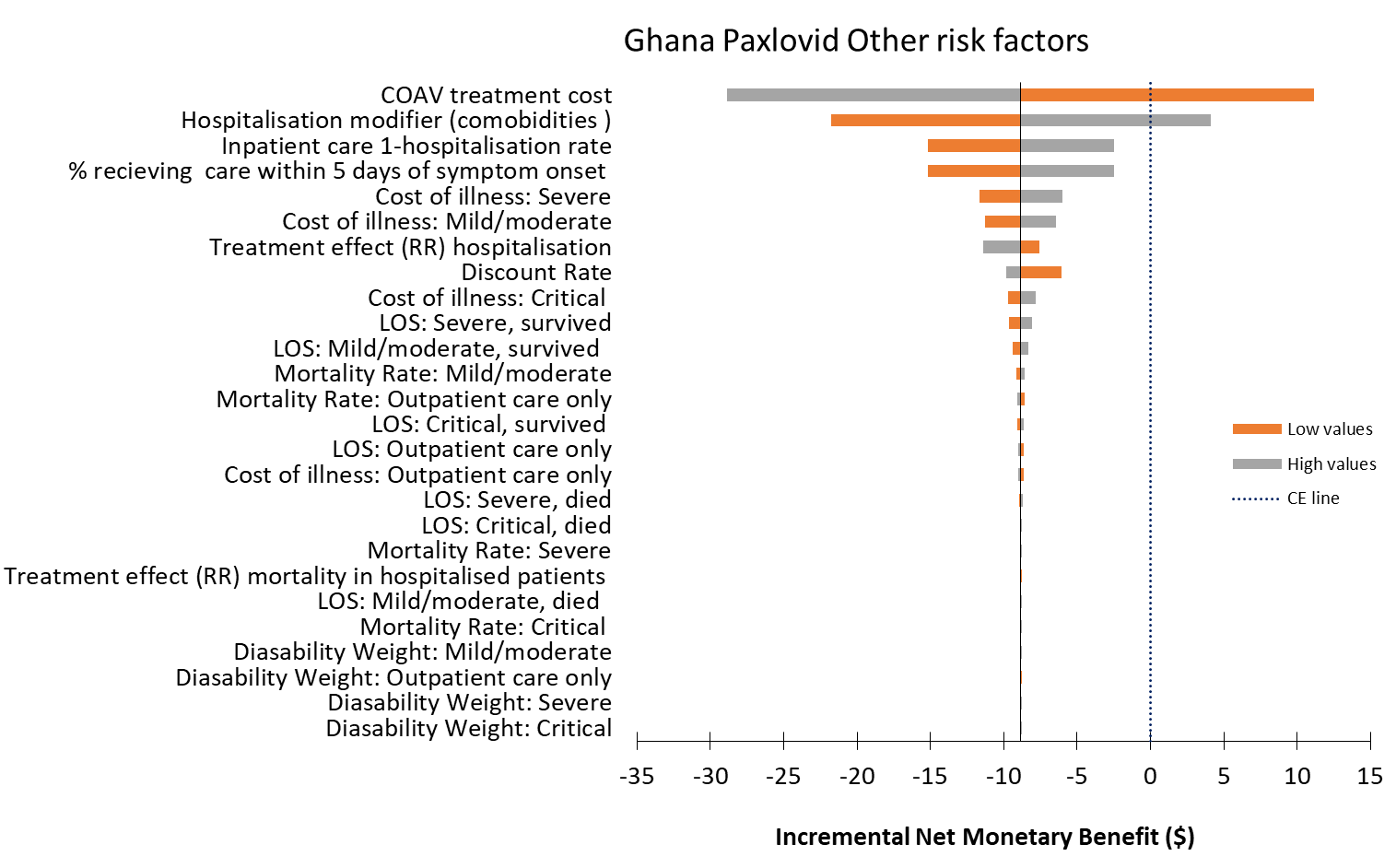

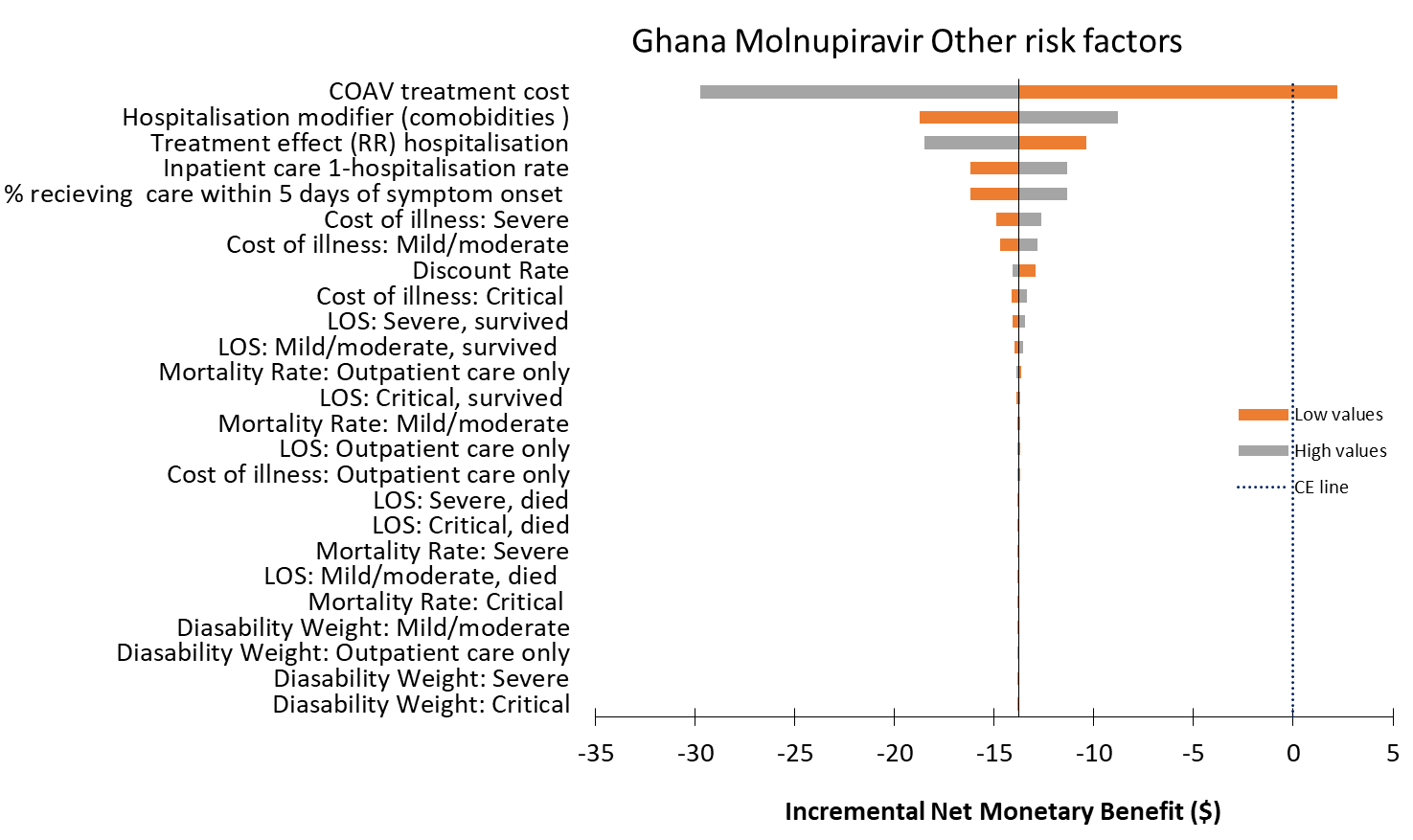

2. **RWANDA**
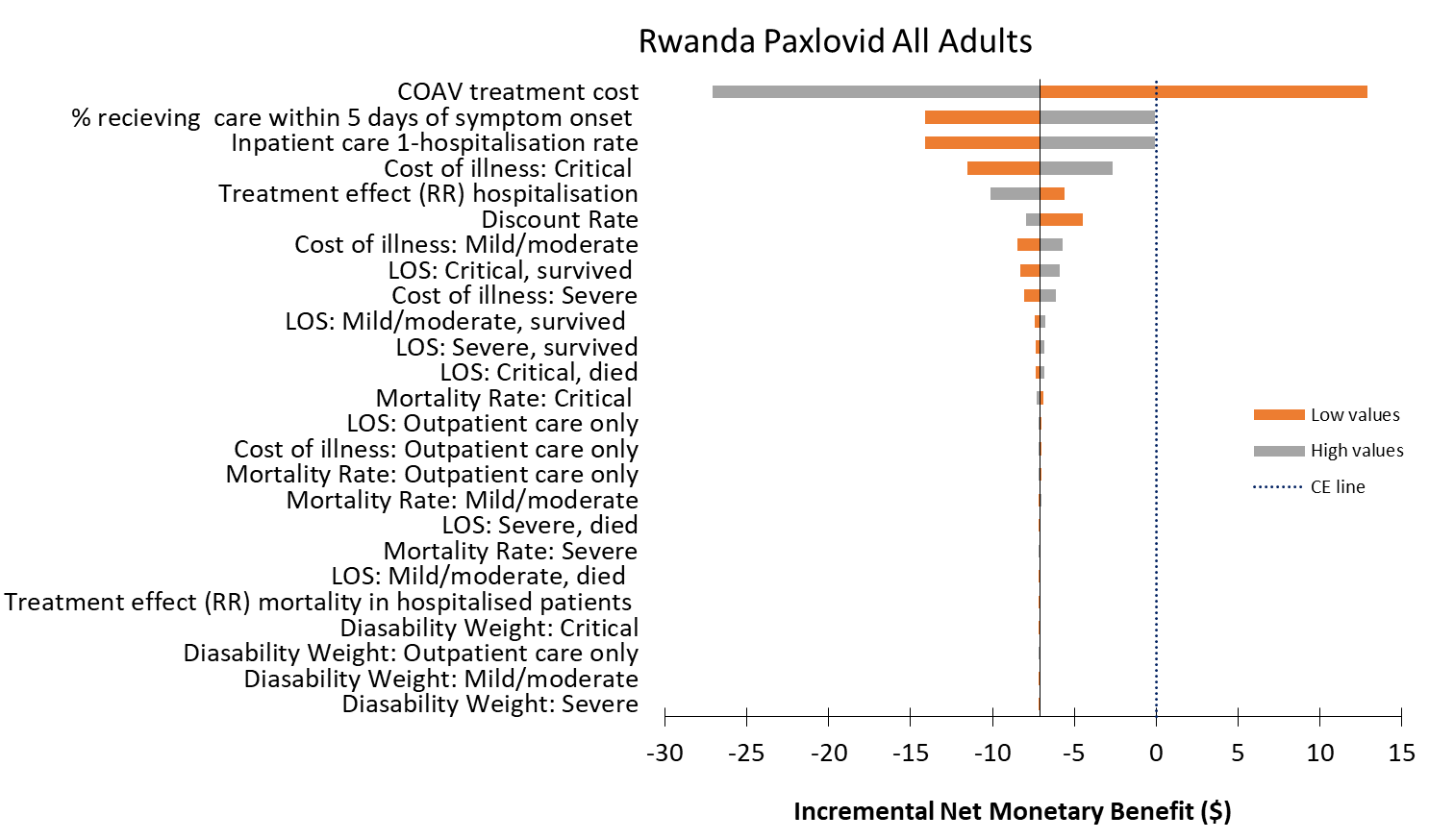

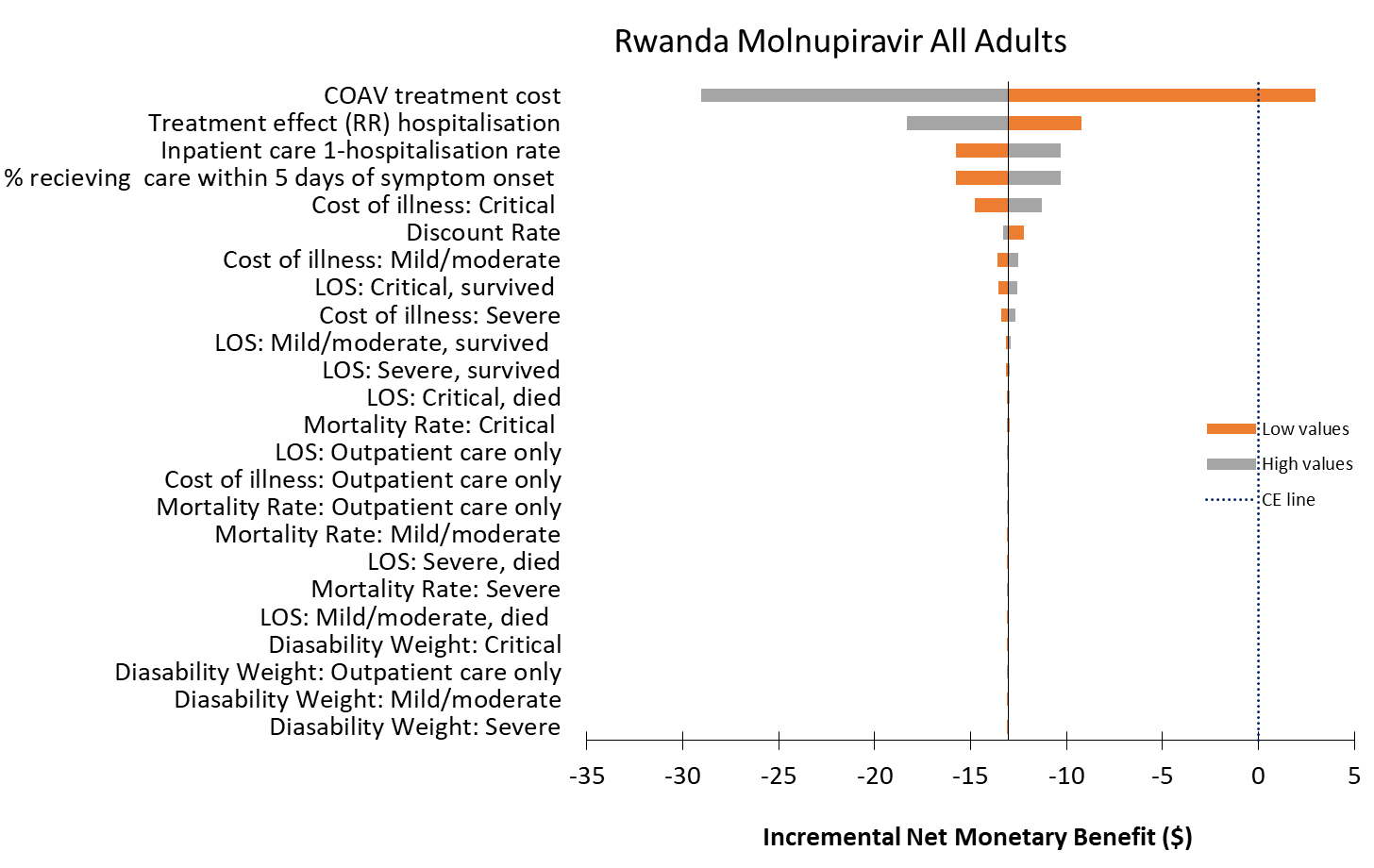

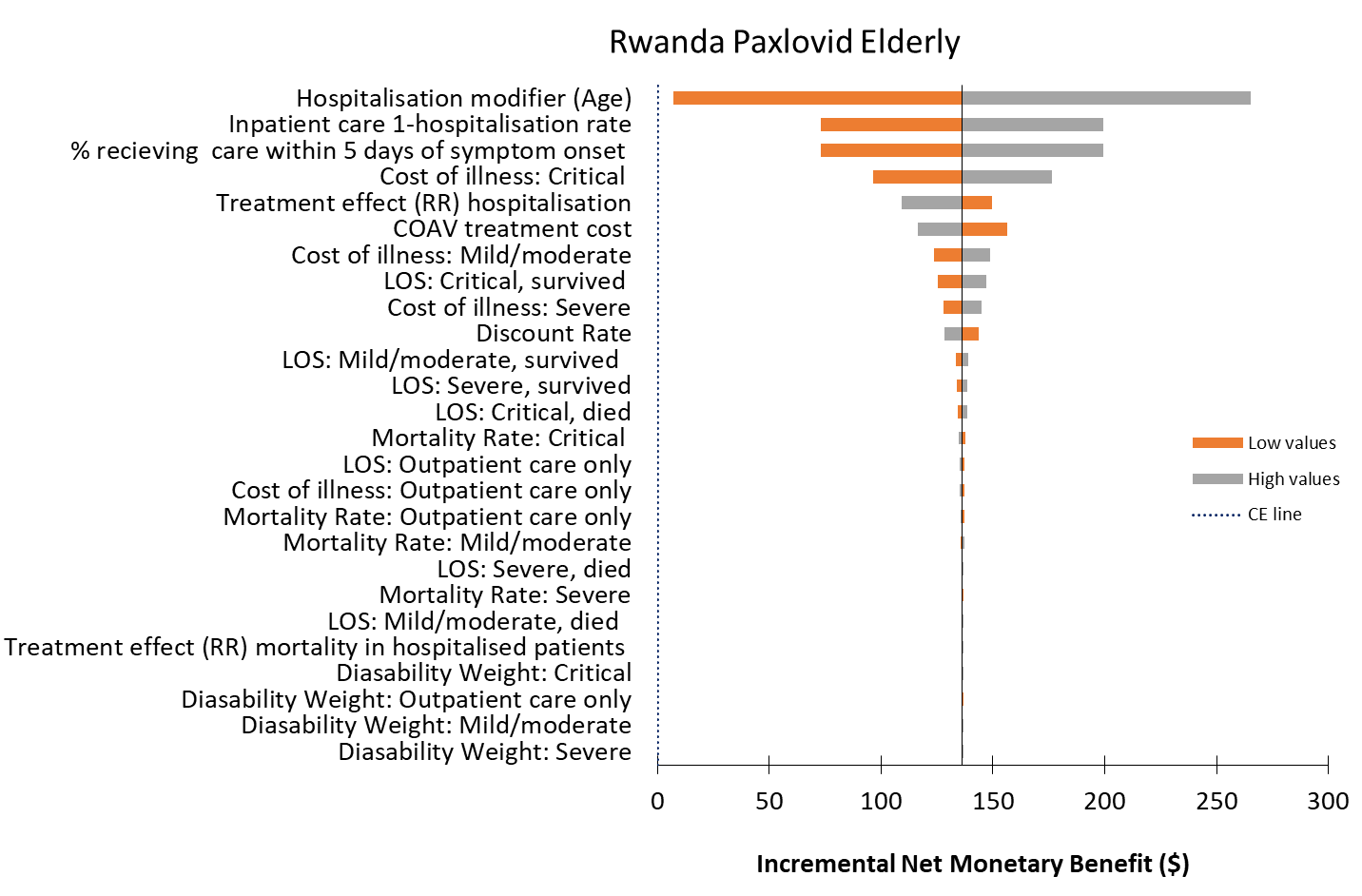

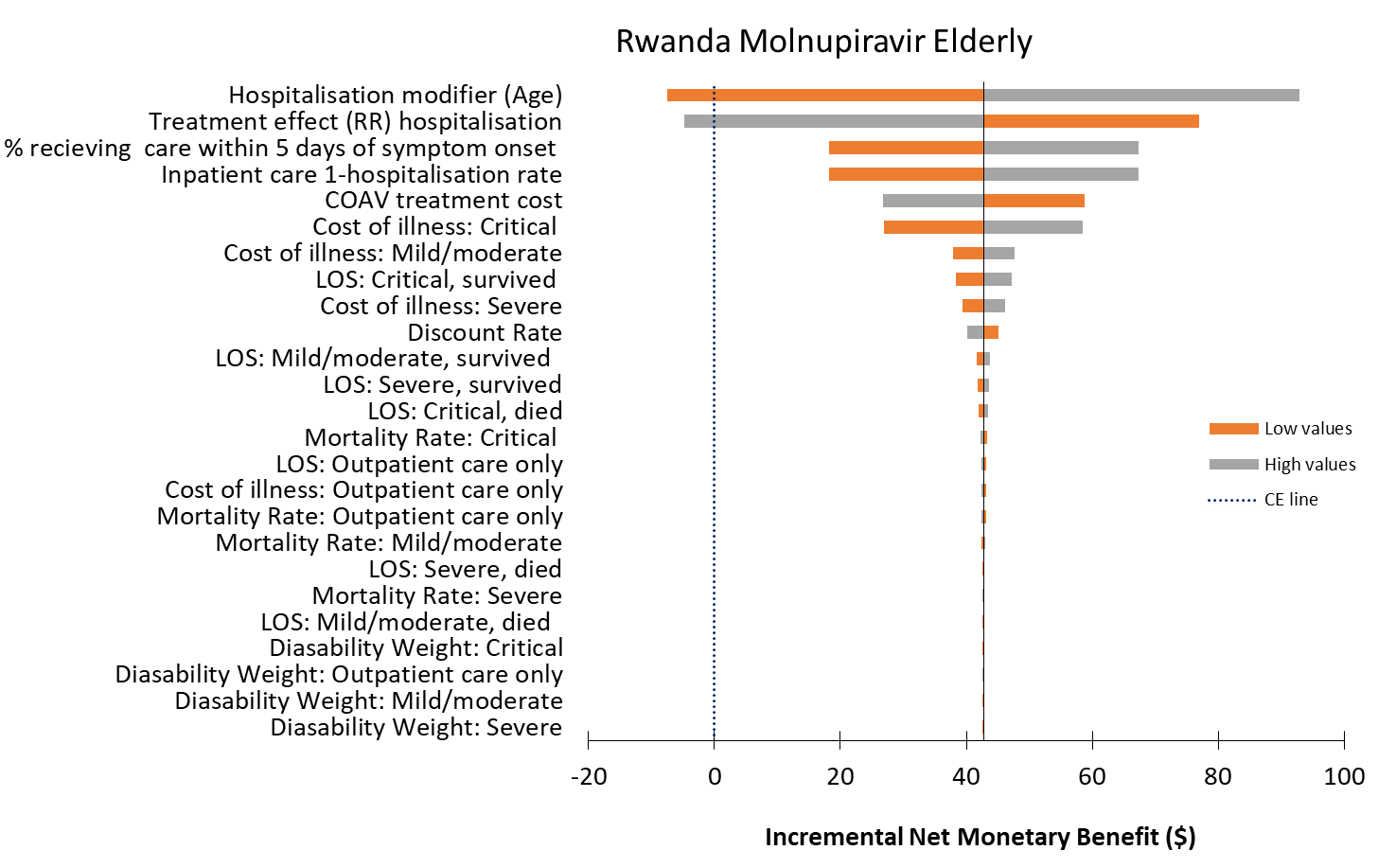

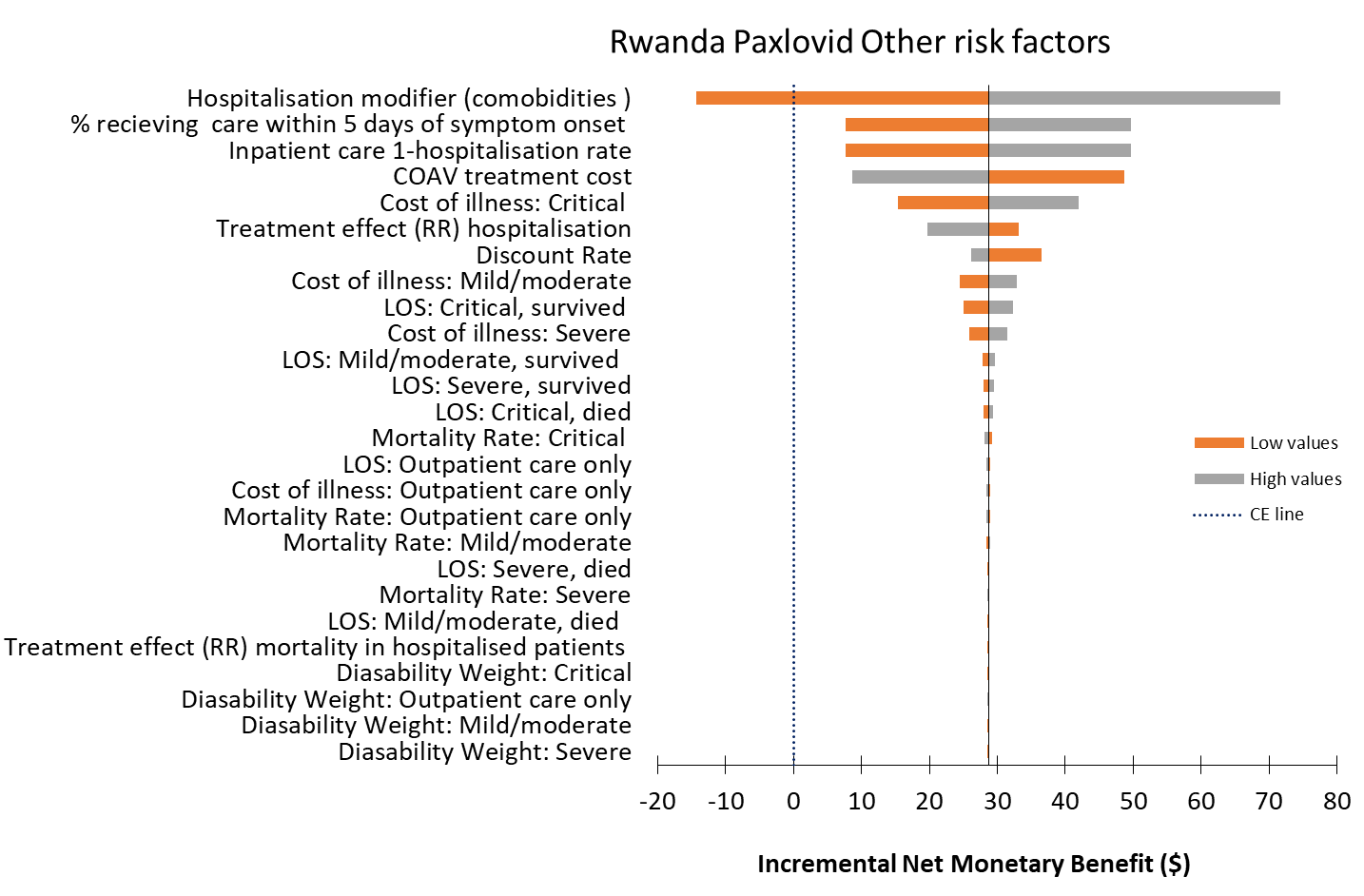

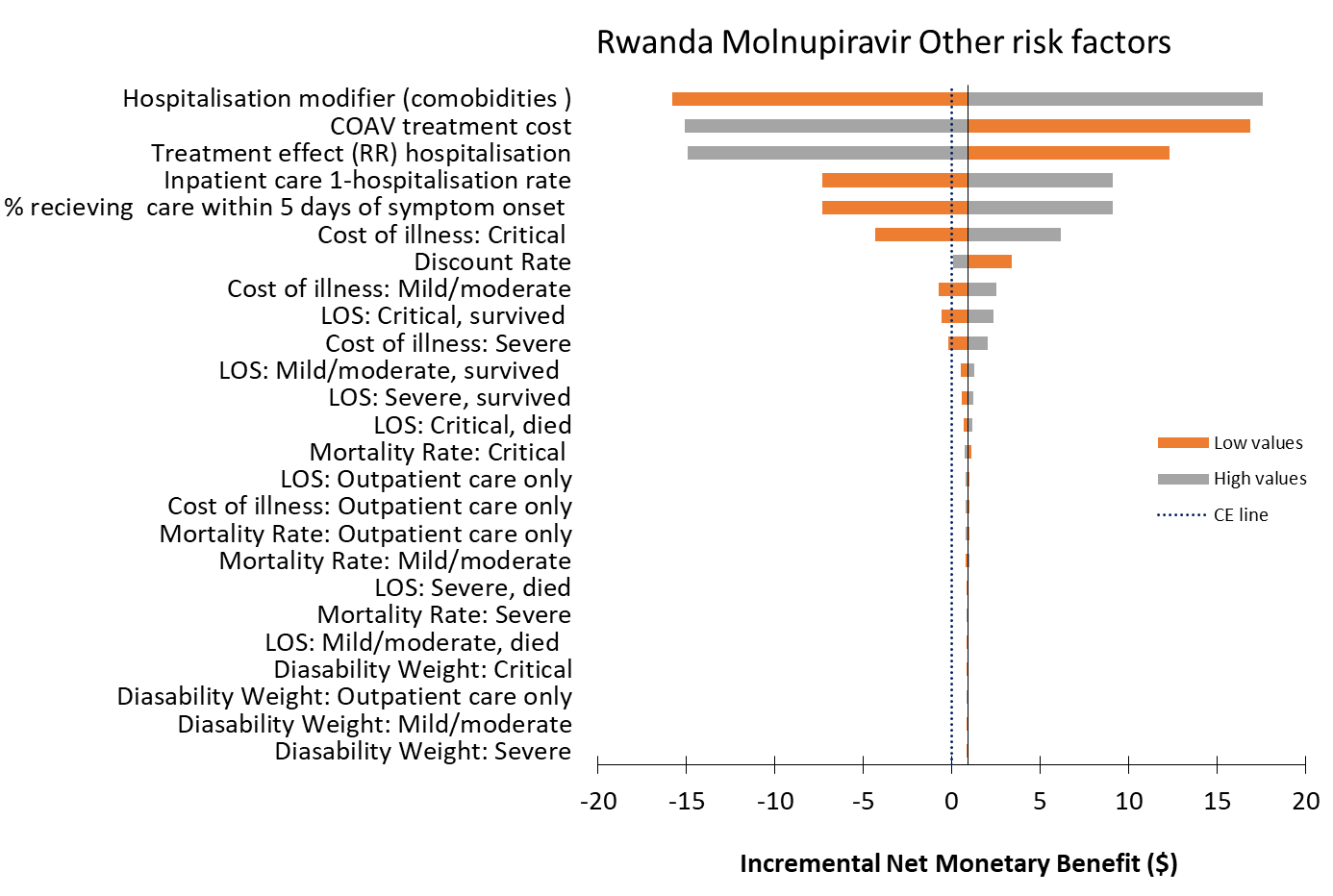

3. **ZAMBIA**
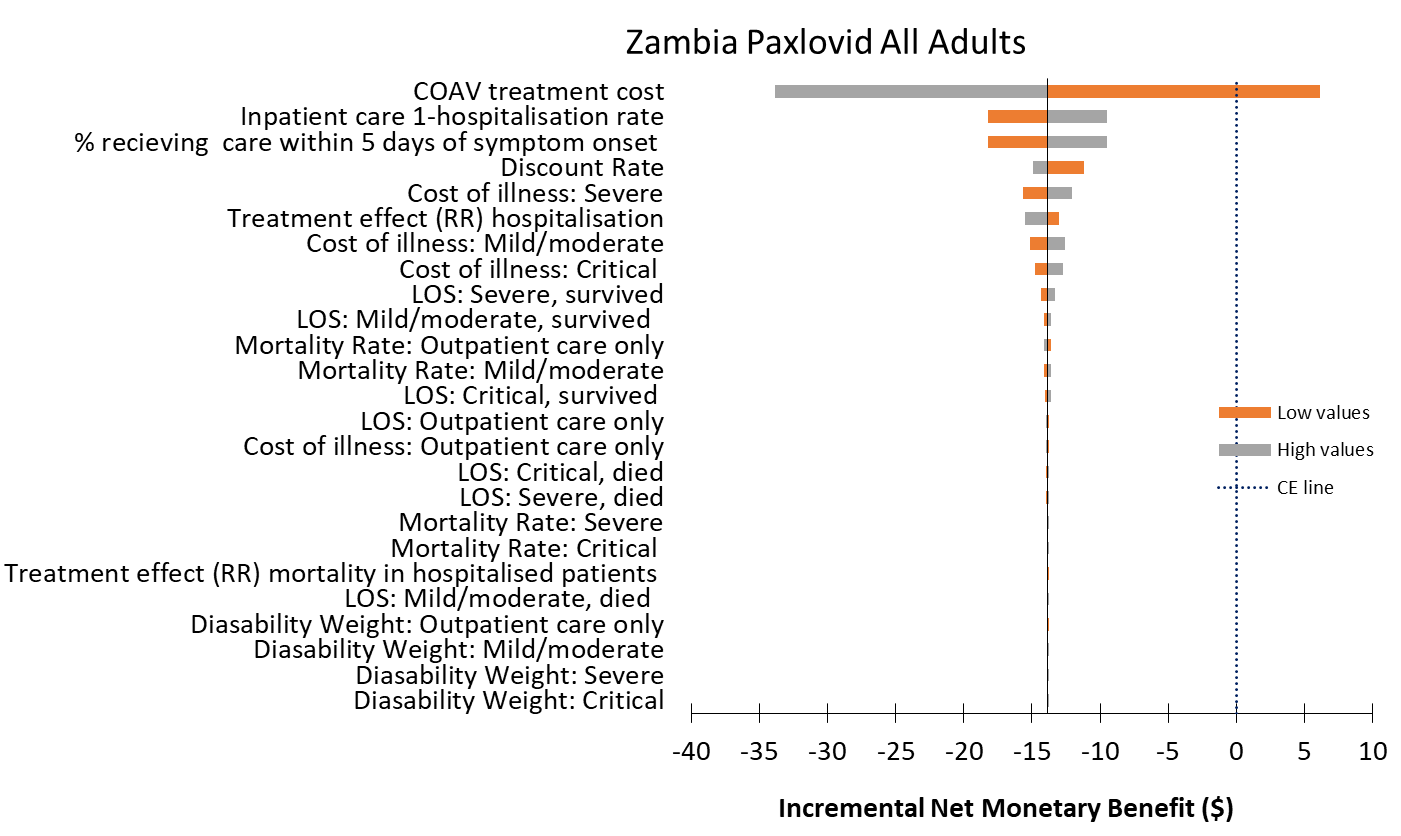

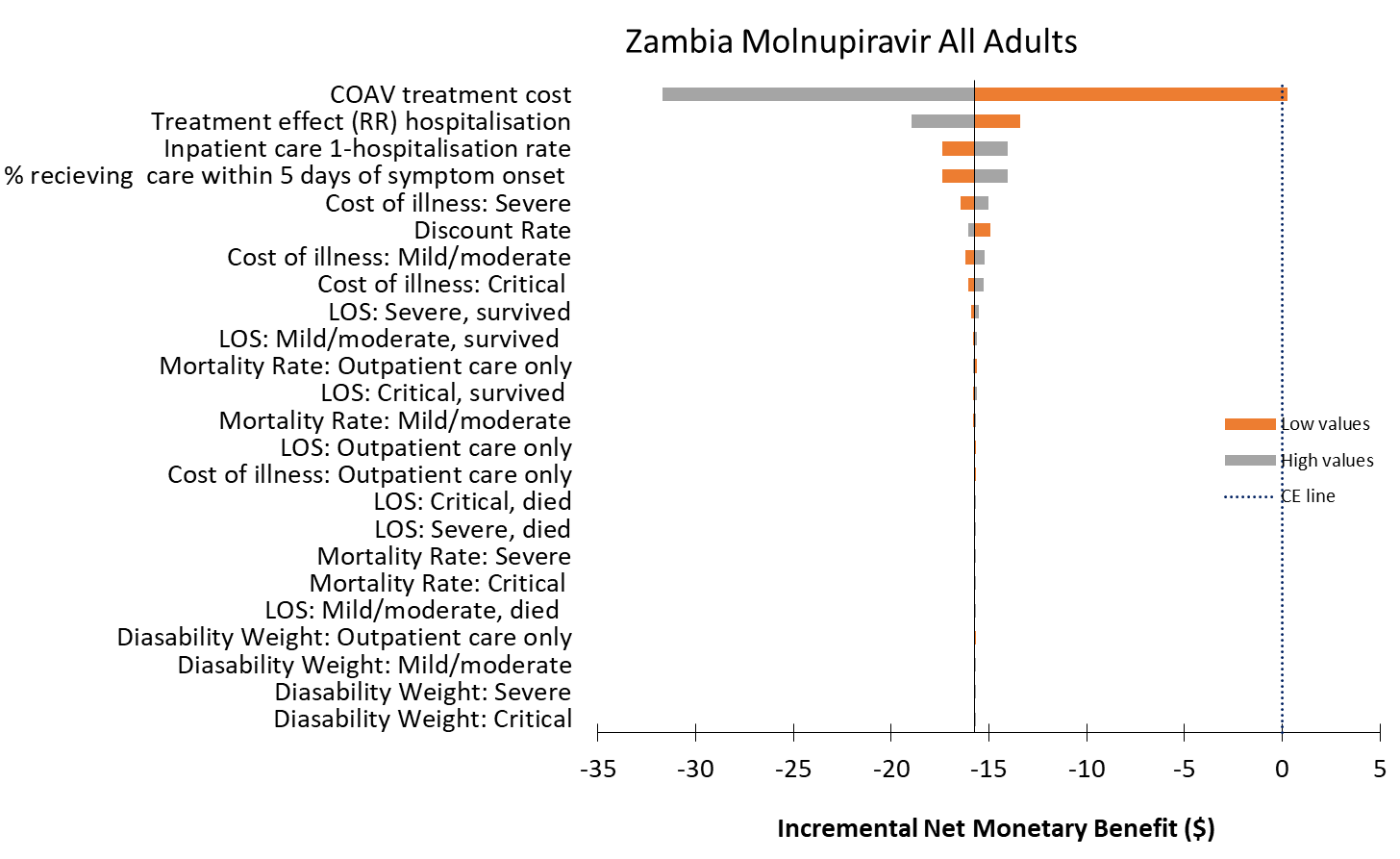

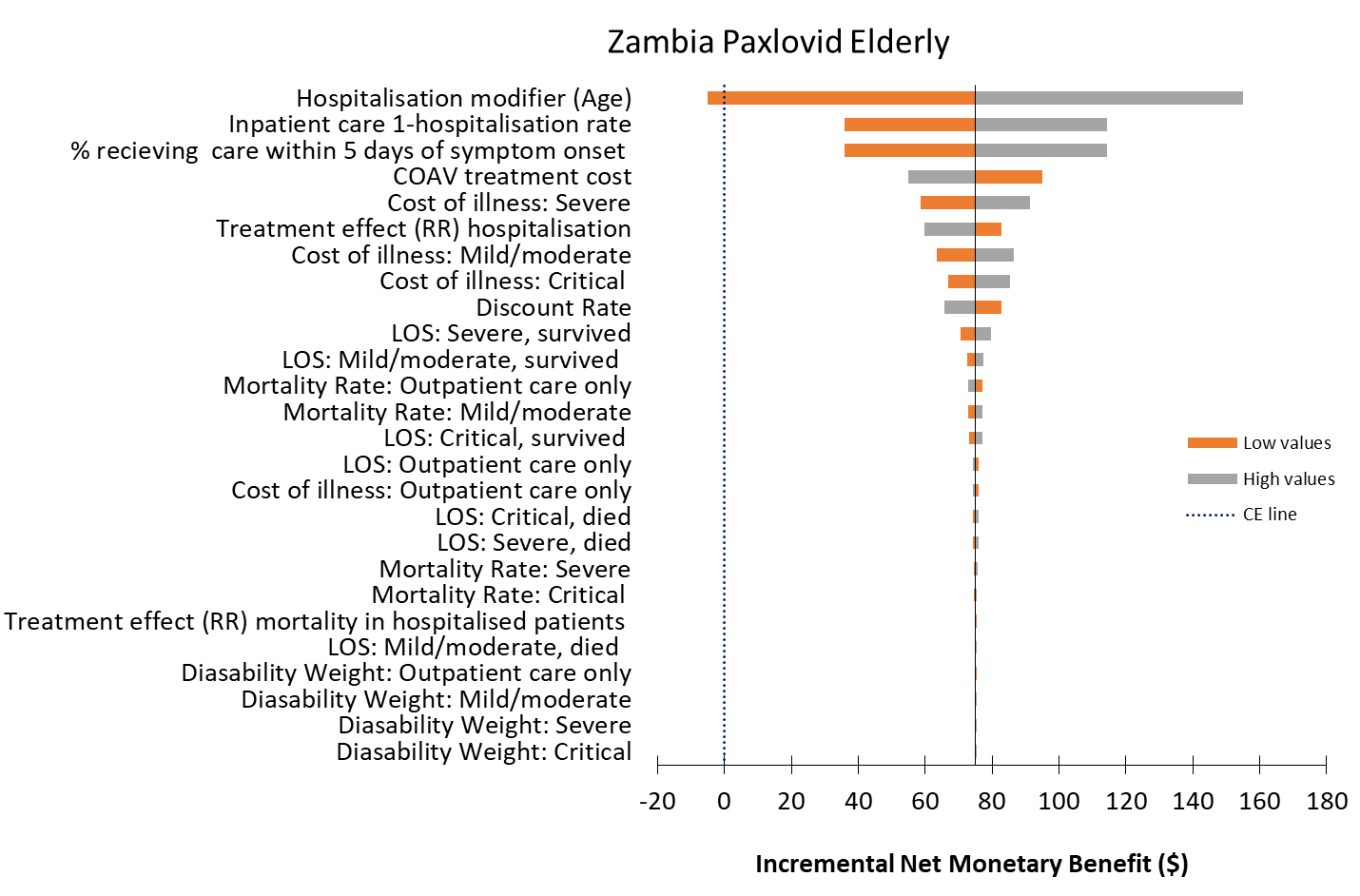

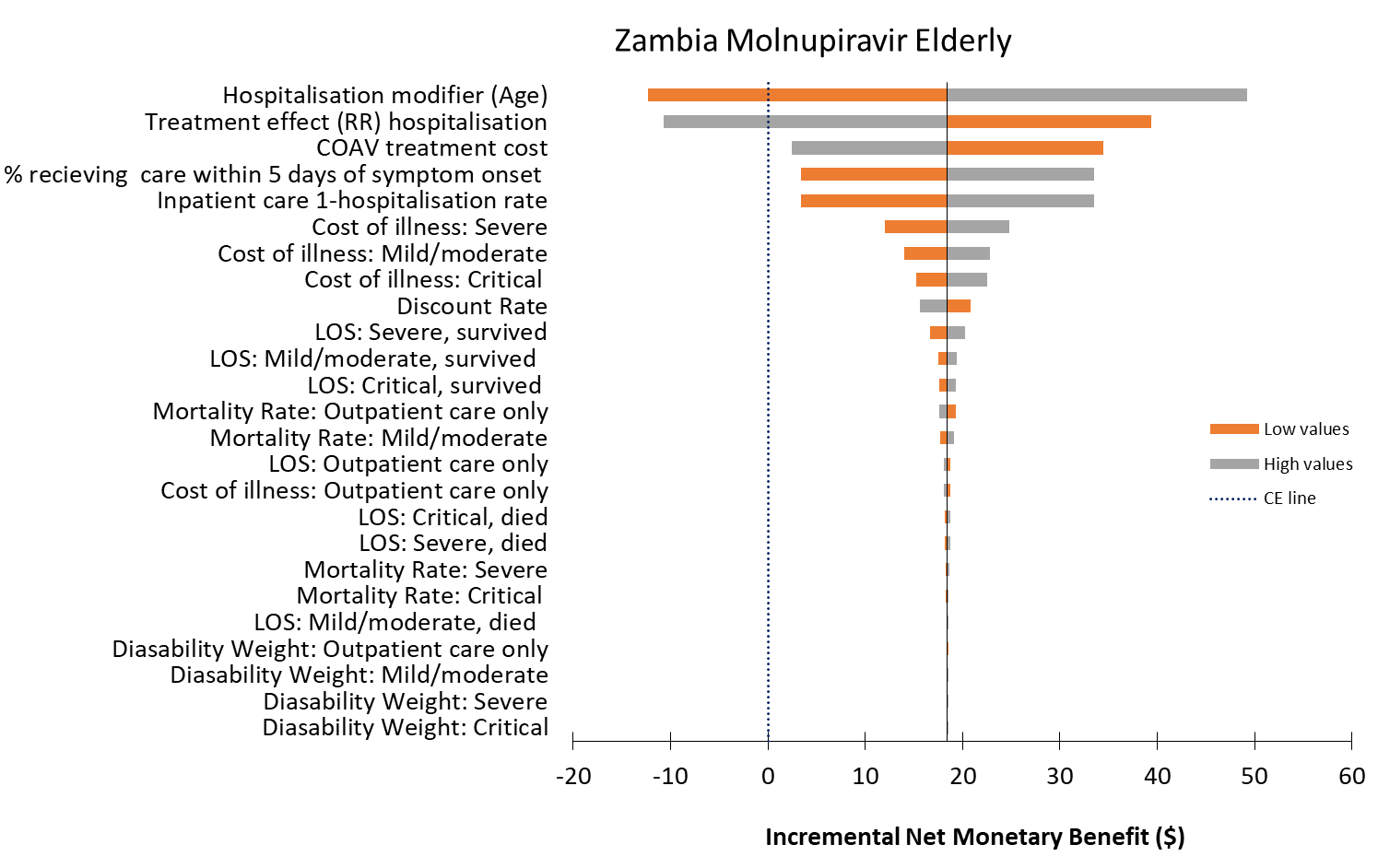

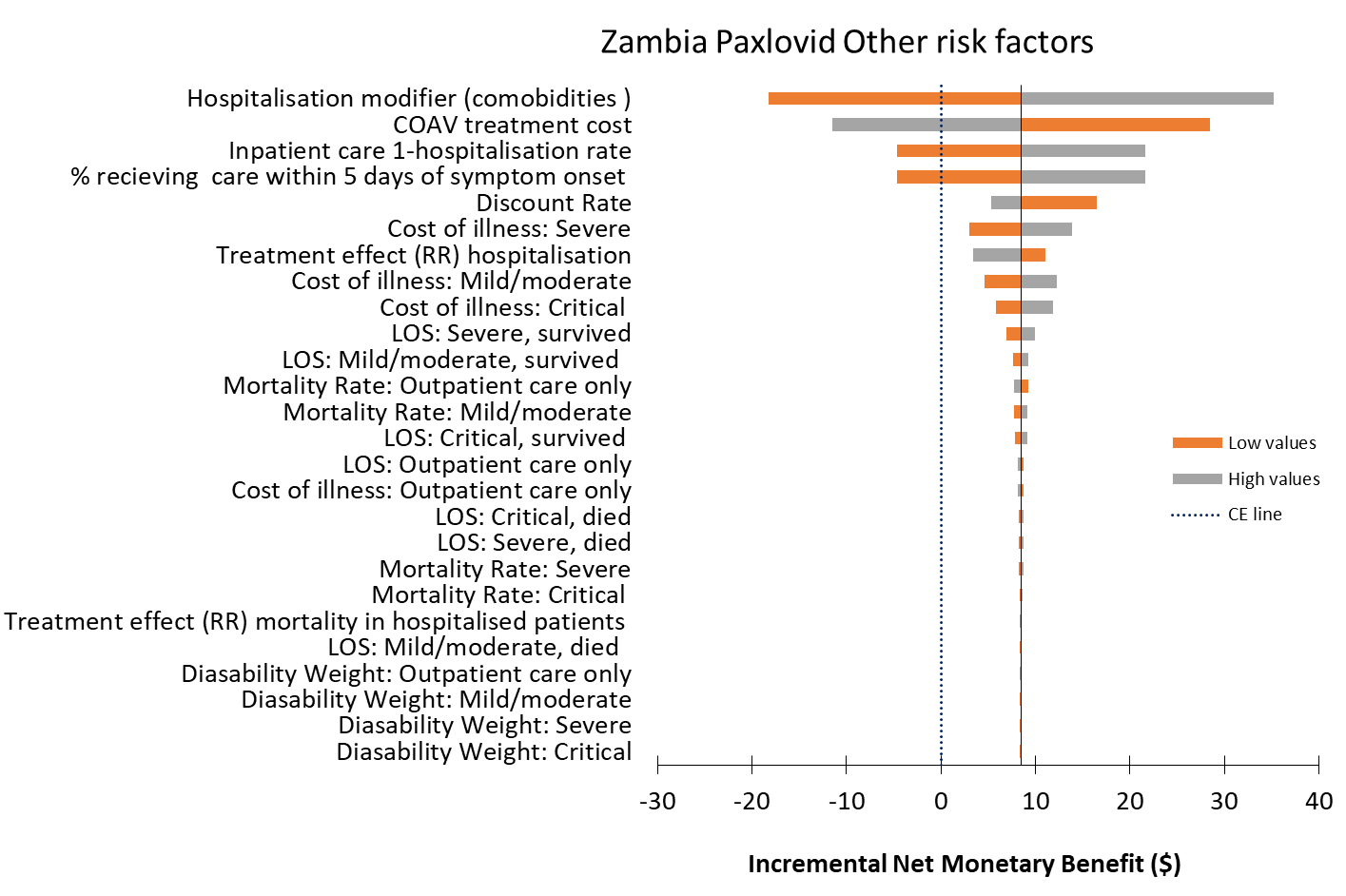

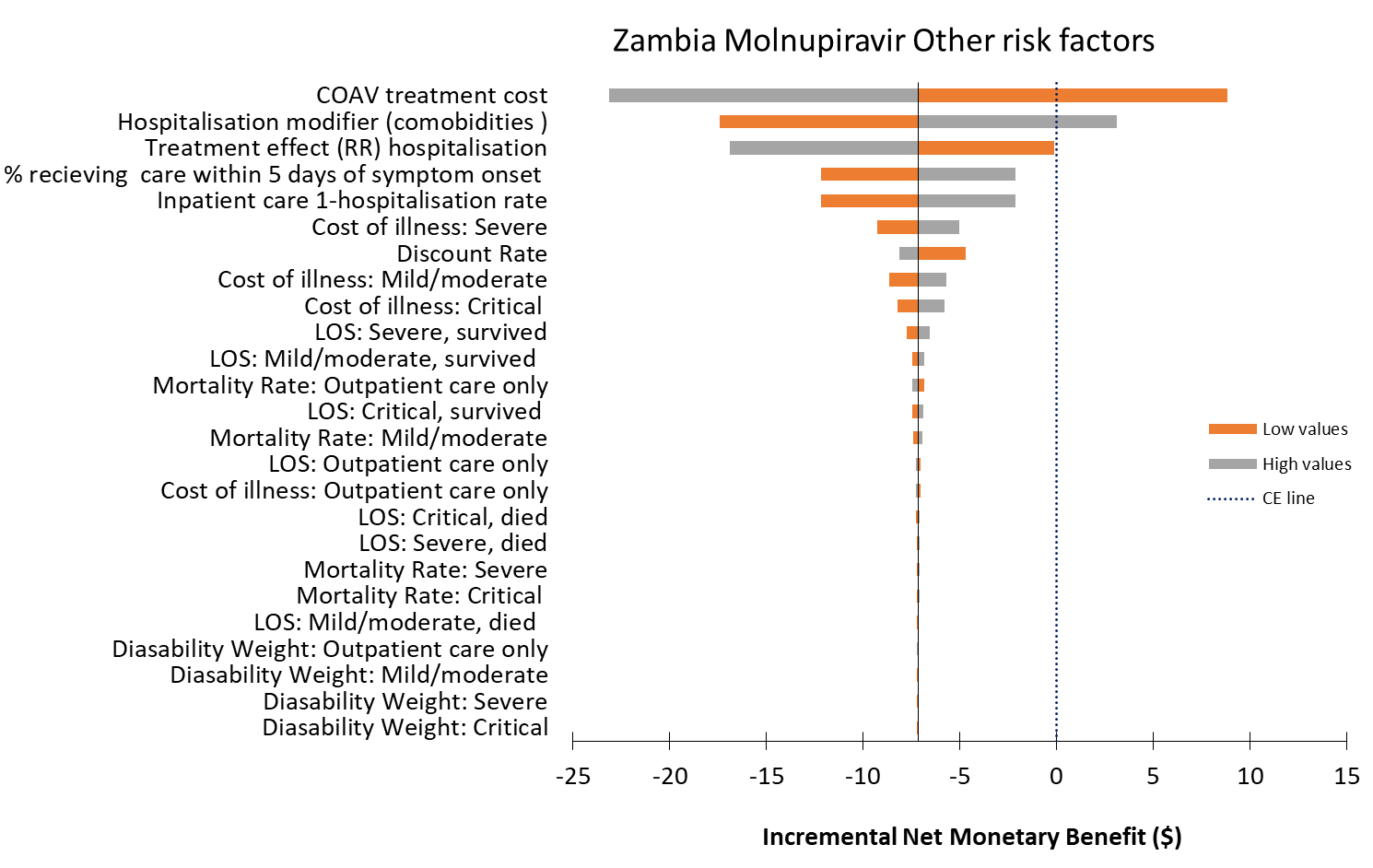

**Table S3:** Incremental outcomes, incremental cost-effectiveness ratios and incremental net monetary benefit (**Scenario Analysis)**

1. ***GHANA***

| ***All adults*** | **Scenario 1** | | **Scenario 2** | | **Scenario 3** | | **Scenario 4** | | **Scenario 5** | |
| --- | --- | --- | --- | --- | --- | --- | --- | --- | --- | --- |
| **Treatment options** | **DALYs** | **Costs (US$)** | **DALYs** | **Costs (US$)** | **DALYs** | **Costs (US$)** | **DALYs** | **Costs (US$)** | **DALYs** | **Costs (US$)** |
| Usual care | 0.2949 | 150.46 | 0.298 | 178.11 | 0.2981 | 784.61 | 0.313 | 202.33 | 0.298 | 901.65 |
| Paxlovid | 0.2945 | 173.69 | 0.288 | 135.21 | 0.2973 | 803.07 | 0.311 | 220.72 | 0.297 | 921.64 |
| Molnupiravir |  |  | 0.295 | 171.57 | 0.2978 | 802.18 | 0.312 | 219.74 | 0.298 | 919.68 |
| **Difference** | **DALYs averted** | **Incr. cost (US$)** | **DALYs averted** | **Incr. cost (US$)** | **DALYs averted** | **Incr. cost (US$)** | **DALYs averted** | **Incr. cost (US$)** | **DALYs averted** | **Incr. cost (US$)** |
| Paxlovid vs Usual care | 0.0004 | 23.22 | 0.010 | -42.90 | 0.0008 | 18.46 | 0.002 | 18.40 | 0.001 | 19.99 |
| Molnupiravir vs Usual care |  |  | 0.003 | -6.54 | 0.0002 | 17.57 | 0.001 | 17.41 | 0.000 | 18.04 |
| **ICER** |  | **ICER** |  | **ICER** |  | **ICER** |  | **ICER** |  | **ICER** |
| Paxlovid vs Usual care |  | 55015.17 |  | -4210.33 |  | 24193.04 |  | 10907.56 |  | 26207.82 |
| Molnupiravir vs Usual care |  |  |  | -2124.29 |  | 76259.90 |  | 29304.05 |  | 78266.13 |
| **INMB** |  | **INMB** |  | **INMB** |  | **INMB** |  | **INMB** |  | **INMB** |
| Paxlovid vs Usual care |  | -23.04 |  | 47.31 |  | -18.13 |  | -17.67 |  | -19.66 |
| Molnupiravir vs Usual care |  |  |  | 7.87 |  | -17.47 |  | -17.16 |  | -17.94 |
| ***Elderly >=65years*** | **Scenario 1** | | **Scenario 2** | | **Scenario 3** | | **Scenario 4** | | **Scenario 5** | |
| **Treatment options** | **DALYs** | **Costs (US$)** | **DALYs** | **Costs (US$)** | **DALYs** | **Costs (US$)** | **DALYs** | **Costs (US$)** | **DALYs** | **Costs (US$)** |
| Usual care | 0.343 | 572.68 | 0.372 | 821.46 | 0.3717 | 848.26 | 0.504 | 1039.43 | 0.372 | 1535.70 |
| Paxlovid | 0.339 | 581.70 | 0.280 | 235.37 | 0.3649 | 827.64 | 0.489 | 1005.00 | 0.365 | 1515.63 |
| Molnupiravir |  |  | 0.344 | 602.62 | 0.3697 | 850.43 | 0.499 | 1036.16 | 0.370 | 1538.02 |
| **Difference** | **DALYs averted** | **Incr. cost (US$)** | **DALYs averted** | **Incr. cost (US$)** | **DALYs averted** | **Incr. cost (US$)** | **DALYs averted** | **Incr. cost (US$)** | **DALYs averted** | **Incr. cost (US$)** |
| Paxlovid vs Usual care | 0.004 | 9.02 | 0.091 | -586.10 | 0.007 | -20.63 | 0.015 | -34.43 | 0.007 | -20.06 |
| Molnupiravir vs Usual care |  |  | 0.028 | -218.85 | 0.002 | 2.17 | 0.005 | -3.27 | 0.002 | 2.33 |
| **ICER** |  | **ICER** |  | **ICER** |  | **ICER** |  | **ICER** |  | **ICER** |
| Paxlovid vs Usual care |  | 2379.56 |  | -6407.45 |  | -3011.64 |  | -2273.58 |  | -2929.75 |
| Molnupiravir vs Usual care |  |  |  | -7920.07 |  | 1047.41 |  | -612.48 |  | 1123.97 |
| **INMB** |  | **INMB** |  | **INMB** |  | **INMB** |  | **INMB** |  | **INMB** |
| Paxlovid vs Usual care |  | -7.38 |  | 625.73 |  | 23.59 |  | 40.99 |  | 23.03 |
| Molnupiravir vs Usual care |  |  |  | 230.82 |  | -1.27 |  | 5.58 |  | -1.43 |
| ***Adults with other risk factors*** | **Scenario 1** | | **Scenario 2** | | **Scenario 3** | | **Scenario 4** | | **Scenario 5** | |
| **Treatment options** | **DALYs** | **Costs (US$)** | **DALYs** | **Costs (US$)** | **DALYs** | **Costs (US$)** | **DALYs** | **Costs (US$)** | **DALYs** | **Costs (US$)** |
| Usual care | 0.307 | 256.02 | 0.317 | 338.95 | 0.317 | 981.00 | 0.361 | 411.60 | 0.317 | 1060.61 |
| Paxlovid | 0.306 | 275.69 | 0.286 | 160.25 | 0.314 | 986.37 | 0.356 | 416.79 | 0.314 | 1070.59 |
| Molnupiravir |  |  | 0.307 | 279.33 | 0.316 | 993.72 | 0.359 | 423.84 | 0.316 | 1074.72 |
| **Difference** | **DALYs averted** | **Incr. cost (US$)** | **DALYs averted** | **Incr. cost (US$)** | **DALYs averted** | **Incr. cost (US$)** | **DALYs averted** | **Incr. cost (US$)** | **DALYs averted** | **Incr. cost (US$)** |
| Paxlovid vs Usual care | 0.001 | 19.67 | 0.031 | -178.70 | 0.002 | 5.37 | 0.005 | 5.19 | 0.002 | 9.98 |
| Molnupiravir vs Usual care |  |  | 0.009 | -59.62 | 0.001 | 12.72 | 0.002 | 12.24 | 0.001 | 14.11 |
| **ICER** |  | **ICER** |  | **ICER** |  | **ICER** |  | **ICER** |  | **ICER** |
| Paxlovid vs Usual care |  | 15533.95 |  | -5846.04 |  | 2345.60 |  | 1025.96 |  | 4360.38 |
| Molnupiravir vs Usual care |  |  |  | -6456.16 |  | 18401.35 |  | 6867.80 |  | 20407.57 |
| **INMB** |  | **INMB** |  | **INMB** |  | **INMB** |  | **INMB** |  | **INMB** |
| Paxlovid vs Usual care |  | -19.12 |  | 191.94 |  | -4.38 |  | -3.00 |  | -8.99 |
| Molnupiravir vs Usual care |  |  |  | 63.62 |  | -12.42 |  | -11.47 |  | -13.81 |

2. ***Rwanda***

| ***All adults*** | **Scenario 1** | | **Scenario 2** | | **Scenario 3** | | **Scenario 4** | | **Scenario 5** | |
| --- | --- | --- | --- | --- | --- | --- | --- | --- | --- | --- |
| **Treatment options** | **DALYs** | **Costs (US$)** | **DALYs** | **Costs (US$)** | **DALYs** | **Costs (US$)** | **DALYs** | **Costs (US$)** | **DALYs** | **Costs (US$)** |
| Usual care | 0.222 | 75.02 | 0.227 | 107.61 | 0.227 | 264.54 | 0.234 | 118.02 | 0.227 | 554.45 |
| Paxlovid | 0.221 | 96.98 | 0.219 | 91.16 | 0.224 | 270.25 | 0.228 | 122.41 | 0.224 | 562.62 |
| Molnupiravir |  |  | 0.225 | 111.34 | 0.226 | 277.13 | 0.232 | 129.92 | 0.226 | 567.83 |
| **Difference** | **DALYs averted** | **Incr. cost (US$)** | **DALYs averted** | **Incr. cost (US$)** | **DALYs averted** | **Incr. cost (US$)** | **DALYs averted** | **Incr. cost (US$)** | **DALYs** | **Costs (US$)** |
| Paxlovid vs Usual care | 0.001 | 21.96 | 0.008 | -16.45 | 0.003 | 5.71 | 0.006 | 4.39 | 0.003 | 8.17 |
| Molnupiravir vs Usual care |  |  | 0.003 | 3.73 | 0.001 | 12.59 | 0.002 | 11.90 | 0.001 | 13.38 |
| **ICER** |  | **ICER** |  | **ICER** |  | **ICER** |  | **ICER** |  | **ICER** |
| Paxlovid vs Usual care |  | 24223.69 |  | -1957.04 |  | 1652.78 |  | 793.32 |  | 2364.96 |
| Molnupiravir vs Usual care |  |  |  | 1382.80 |  | 11351.42 |  | 6154.86 |  | 12062.32 |
| **INMB** |  | **INMB** |  | **INMB** |  | **INMB** |  | **INMB** |  | **INMB** |
| Paxlovid vs Usual care |  | -21.73 |  | 18.52 |  | -4.86 |  | -3.03 |  | -7.32 |
| Molnupiravir vs Usual care |  |  |  | -3.07 |  | -12.32 |  | -11.42 |  | -13.11 |
| ***Elderly >=65years*** | **Scenario 1** | | **Scenario 2** | | **Scenario 3** | | **Scenario 4** | | **Scenario 5** | |
| **Treatment options** | **DALYs** | **Costs (US$)** | **DALYs** | **Costs (US$)** | **DALYs** | **Costs (US$)** | **DALYs** | **Costs (US$)** | **DALYs** | **Costs (US$)** |
| Usual care | 0.251 | 208.74 | 0.301 | 502.02 | 0.301 | 510.15 | 0.358 | 595.68 | 0.301 | 957.63 |
| Paxlovid | 0.243 | 206.36 | 0.224 | 153.95 | 0.269 | 381.84 | 0.306 | 435.21 | 0.269 | 831.25 |
| Molnupiravir |  |  | 0.276 | 375.59 | 0.291 | 469.98 | 0.340 | 542.77 | 0.291 | 918.07 |
| **Difference** | **DALYs averted** | **Incr. cost (US$)** | **DALYs averted** | **Incr. cost (US$)** | **DALYs averted** | **Incr. cost (US$)** | **DALYs averted** | **Incr. cost (US$)** | **DALYs** | **Costs (US$)** |
| Paxlovid vs Usual care | 0.008 | -2.38 | 0.078 | -348.06 | 0.032 | -128.30 | 0.051 | -160.47 | 0.032 | -126.38 |
| Molnupiravir vs Usual care |  |  | 0.025 | -126.42 | 0.010 | -40.17 | 0.018 | -52.91 | 0.010 | -39.56 |
| **ICER** |  | **ICER** |  | **ICER** |  | **ICER** |  | **ICER** |  | **ICER** |
| Paxlovid vs Usual care |  | -283.47 |  | -4471.10 |  | -4008.59 |  | -3130.55 |  | -3948.42 |
| Molnupiravir vs Usual care |  |  |  | -5060.19 |  | -3910.64 |  | -2956.25 |  | -3851.01 |
| **INMB** |  | **INMB** |  | **INMB** |  | **INMB** |  | **INMB** |  | **INMB** |
| Paxlovid vs Usual care |  | 4.45 |  | 367.25 |  | 136.19 |  | 173.11 |  | 134.27 |
| Molnupiravir vs Usual care |  |  |  | 132.58 |  | 42.70 |  | 57.32 |  | 42.09 |
| ***Adults with other risk factors*** | **Scenario 1** | | **Scenario 2** | | **Scenario 3** | | **Scenario 4** | | **Scenario 5** | |
| **Treatment options** | **DALYs** | **Costs (US$)** | **DALYs** | **Costs (US$)** | **DALYs** | **Costs (US$)** | **DALYs** | **Costs (US$)** | **DALYs** | **Costs (US$)** |
| Usual care | 0.228 | 108.45 | 0.244 | 206.21 | 0.244 | 373.74 | 0.262 | 237.43 | 0.244 | 652.02 |
| Paxlovid | 0.225 | 124.33 | 0.219 | 106.86 | 0.233 | 340.88 | 0.245 | 200.61 | 0.233 | 626.55 |
| Molnupiravir |  |  | 0.236 | 177.40 | 0.241 | 371.52 | 0.256 | 233.13 | 0.241 | 652.17 |
| **Difference** | **DALYs averted** | **Incr. cost (US$)** | **DALYs averted** | **Incr. cost (US$)** | **DALYs averted** | **Incr. cost (US$)** | **DALYs averted** | **Incr. cost (US$)** | **DALYs** | **Costs (US$)** |
| Paxlovid vs Usual care | 0.003 | 15.87 | 0.025 | -99.35 | 0.010 | -32.86 | 0.017 | -36.82 | 0.010 | -25.48 |
| Molnupiravir vs Usual care |  |  | 0.008 | -28.81 | 0.003 | -2.22 | 0.006 | -4.30 | 0.003 | 0.14 |
| **ICER ( US$)** |  | **ICER** |  | **ICER** |  | **ICER** |  | **ICER** |  | **ICER** |
| Paxlovid vs Usual care |  | 5837.15 |  | -3939.65 |  | -3169.34 |  | -2216.97 |  | -2457.16 |
| Molnupiravir vs Usual care |  |  |  | -3559.18 |  | -668.47 |  | -741.99 |  | 42.44 |
| **INMB (US$)** |  | **INMB** |  | **INMB** |  | **INMB** |  | **INMB** |  | **INMB** |
| Paxlovid vs Usual care |  | -15.20 |  | 105.57 |  | 35.42 |  | 40.92 |  | 28.03 |
| Molnupiravir vs Usual care |  |  |  | 30.80 |  | 3.04 |  | 5.73 |  | 0.68 |

3. ***Zambia***

| ***All adults*** | **Scenario 1** | | **Scenario 2** | | **Scenario 3** | | **Scenario 4** | | **Scenario 5** | |
| --- | --- | --- | --- | --- | --- | --- | --- | --- | --- | --- |
| **Treatment options** | **DALYs** | **Costs (US$)** | **DALYs** | **Costs (US$)** | **DALYs** | **Costs (US$)** | **DALYs** | **Costs (US$)** | **Expected DALYs** | **Expected costs (US$)** |
| Usual care | 0.428 | 140.82 | 0.429 | 149.38 | 0.429 | 519.51 | 0.442 | 166.42 | 0.429 | 893.37 |
| Paxlovid | 0.426 | 161.12 | 0.417 | 117.76 | 0.427 | 532.54 | 0.437 | 178.67 | 0.427 | 908.39 |
| Molnupiravir |  |  | 0.425 | 147.17 | 0.428 | 534.97 | 0.440 | 181.40 | 0.428 | 909.44 |
| **Difference** | **DALYs averted** | **Incr. cost (US$)** | **DALYs averted** | **Incr. cost (US$)** | **DALYs averted** | **Incr. cost (US$)** | **DALYs averted** | **Incr. cost (US$)** | **DALYs averted** | **Incr. cost (US$)** |
| Paxlovid vs Usual care | 0.002 | 20.30 | 0.012 | -31.61 | 0.002 | 13.04 | 0.004 | 12.25 | 0.002 | 15.02 |
| Molnupiravir vs Usual care |  |  | 0.004 | -2.21 | 0.001 | 15.46 | 0.001 | 14.98 | 0.001 | 16.06 |
| **ICER** |  | **ICER** |  | **ICER (US$)** |  | **ICER (US$)** |  | **ICER (US$)** |  | **ICER (US$)** |
| Paxlovid vs Usual care |  | 13016.21 |  | -2673.42 |  | 6117.25 |  | 3042.43 |  | 7047.05 |
| Molnupiravir vs Usual care |  |  |  | -612.95 |  | 23789.62 |  | 10686.89 |  | 24717.06 |
| **INMB (US$)** |  | **INMB** |  | **INMB** |  | **INMB** |  | **INMB** |  | **INMB** |
| Paxlovid vs Usual care |  | -19.52 |  | 37.57 |  | -11.96 |  | -10.22 |  | -13.94 |
| Molnupiravir vs Usual care |  |  |  | 4.03 |  | -15.13 |  | -14.28 |  | -15.74 |
| ***Elderly >=65years*** | **Scenario 1** | | **Scenario 2** | | **Scenario 3** | | **Scenario 4** | | **Scenario 5** | |
| **Treatment options** | **DALYs** | **Costs (US$)** | **DALYs** | **Costs (US$)** | **DALYs** | **Costs (US$)** | **DALYs** | **Costs (US$)** | **Expected DALYs** | **Expected costs (US$)** |
| Usual care | 0.487 | 610.79 | 0.498 | 687.82 | 0.498 | 699.88 | 0.608 | 841.20 | 0.498 | 1396.85 |
| Paxlovid | 0.473 | 593.53 | 0.395 | 203.30 | 0.480 | 633.15 | 0.573 | 751.41 | 0.480 | 1331.93 |
| Molnupiravir |  |  | 0.467 | 507.92 | 0.492 | 683.89 | 0.596 | 816.05 | 0.492 | 1381.40 |
| **Difference** | **DALYs averted** | **Incr. cost (US$)** | **DALYs averted** | **Incr. cost (US$)** | **DALYs averted** | **Incr. cost (US$)** | **DALYs averted** | **Incr. cost (US$)** | **DALYs averted** | **Incr. cost (US$)** |
| Paxlovid vs Usual care | 0.014 | -17.27 | 0.103 | -484.52 | 0.019 | -66.73 | 0.035 | -89.79 | 0.019 | -64.91 |
| Molnupiravir vs Usual care |  |  | 0.031 | -179.90 | 0.006 | -15.99 | 0.012 | -25.14 | 0.006 | -15.44 |
| **ICER** |  | **ICER** |  | **ICER (US$)** |  | **ICER (US$)** |  | **ICER (US$)** |  | **ICER (US$)** |
| Paxlovid vs Usual care |  | -1274.98 |  | -4718.71 |  | -3606.21 |  | -2568.40 |  | -3508.14 |
| Molnupiravir vs Usual care |  |  |  | -5744.19 |  | -2832.91 |  | -2064.66 |  | -2736.22 |
| **INMB (US$)** |  | **INMB** |  | **INMB** |  | **INMB** |  | **INMB** |  | **INMB** |
| Paxlovid vs Usual care |  | 24.09 |  | 536.22 |  | 76.05 |  | 107.39 |  | 74.23 |
| Molnupiravir vs Usual care |  |  |  | 195.66 |  | 18.83 |  | 31.28 |  | 18.28 |
| ***Adults with comorbidities*** | **Scenario 1** | | **Scenario 2** | | **Scenario 3** | | **Scenario 4** | | **Scenario 5** | |
| **Treatment options** | **DALYs** | **Costs (US$)** | **DALYs** | **Costs (US$)** | **DALYs** | **Costs (US$)** | **DALYs** | **Costs (US$)** | **Expected DALYs** | **Expected costs (US$)** |
| Usual care | 0.447 | 258.31 | 0.451 | 283.99 | 0.451 | 672.14 | 0.489 | 335.11 | 0.451 | 1025.73 |
| Paxlovid | 0.442 | 269.22 | 0.415 | 139.15 | 0.444 | 661.25 | 0.477 | 321.85 | 0.444 | 1020.79 |
| Molnupiravir |  |  | 0.440 | 237.36 | 0.449 | 678.52 | 0.484 | 340.07 | 0.449 | 1033.93 |
| **Difference** | **DALYs averted** | **Incr. cost (US$)** | **DALYs averted** | **Incr. cost (US$)** | **DALYs averted** | **Incr. cost (US$)** | **DALYs averted** | **Incr. cost (US$)** | **DALYs averted** | **Incr. cost (US$)** |
| Paxlovid vs Usual care | 0.005 | 10.91 | 0.035 | -144.84 | 0.006 | -10.89 | 0.012 | -13.26 | 0.006 | -4.95 |
| Molnupiravir vs Usual care |  |  | 0.011 | -46.63 | 0.002 | 6.38 | 0.004 | 4.95 | 0.002 | 8.19 |
| **ICER** |  | **ICER** |  | **ICER (US$)** |  | **ICER (US$)** |  | **ICER (US$)** |  | **ICER (US$)** |
| Paxlovid vs Usual care |  | 2331.52 |  | -4082.86 |  | -1703.84 |  | -1098.39 |  | -774.03 |
| Molnupiravir vs Usual care |  |  |  | -4309.98 |  | 3274.51 |  | 1177.25 |  | 4201.94 |
| **INMB (US$)** |  | **INMB** |  | **INMB** |  | **INMB** |  | **INMB** |  | **INMB** |
| Paxlovid vs Usual care |  | -8.55 |  | 162.70 |  | 14.11 |  | 19.34 |  | 8.17 |
| Molnupiravir vs Usual care |  |  |  | 52.08 |  | -5.40 |  | -2.83 |  | -7.21 |
